# Contribution of prenatal exposure to ambient temperature extremes and severe maternal morbidity: A retrospective Southern birth cohort

**DOI:** 10.1101/2022.06.11.22276277

**Authors:** Jennifer D. Runkle, Maggie M. Sugg, Scott E. Stevens

**Author notes:** corresponding authorEmail.

## Abstract

**BACKGROUND:** Health disparities have persisted in severe maternal morbidity (SMM), an event in which a woman nearly dies from a complication during pregnancy, with limited data on environmental risk factors.

****OBJECTIVE**:** To examine the association between prenatal exposure to high and low ambient temperatures and SMM during critical windows of pregnancy for a birth cohort in the Southeastern United States.

**METHODS:** This retrospective, population-based birth cohort included hospital deliveries from 1999 to 2017 (570,660 women, 921,444 deliveries). Daily average temperatures at the county-scale were merged with delivery discharge records and days of exposure to very hot and very cold were estimated over the following critical windows: preconception, and first, second, and third trimesters (T1-T3). Generalized estimating equations with multivariable Poisson models examined the association between temperature extremes and SMM for each critical window.

**RESULTS:** Women exposed to a low compared to a high number of cold days during the first and third trimesters were 1.11 (CI: 1.03, 1.20) and 1.30 (CI: 1.20, 1.42) times more likely to experience SMM, respectively. Compared to the no exposure group, women exposed to a high number of very hot temperatures during preconception were 1.09 (95%CI:1.02,1.18) more likely to experience SMM. Sustained exposure to a high or moderate-intensity heat wave during the summer months was associated with a 45% or 39% increase in SMM risk during T2, respectively. Pregnant populations residing in rural locations were more sensitive to cold exposure in T3. Women exposed to a high number of very hot days in T2 compared to no exposure were 20% more likely to experience preterm SMM.

****SIGNIFICANCE**:** Findings suggest that maternal exposure to hot or cold temperature extremes around the time or during pregnancy may be a contributing environmental risk factor for SMM. More attention should be focused on prenatal counseling in pregnant populations around the risk of thermal extremes.

**Impact Statement:** This is the first study to examine the association between severe maternal morbidity and ambient cold and hot temperature extremes. Results revealed an increase in SMM risk for pregnant individuals following unseasonably cold exposure during the first and third trimesters and exposure to hotter than average temperatures in the second trimester. Our findings suggest that maternal exposure to ambient temperature extremes is a modifiable risk factor for SMM. This study considered contextual social and environmental factors associated with increased SMM risks, such as residential segregation (a proxy for structural racism), residential poverty, and rural compared to urban differences.

## Introduction

Emerging research demonstrates that everyday environmental exposures, like hot ambient temperature, are being amplified in the presence of a changing climate resulting in adverse maternal health outcomes (1, 2). One mechanism by which climate change may impact pregnancy health is through prolonged exposure to thermal extremes (3, 4). An important scientific gap remains in understanding the impact of cold and hot ambient temperatures and pregnancy risks in a changing climate, particularly for Black birthing populations.

For every woman who dies from complications in pregnancy, another 50 to 100 women experience a “near miss (mortality)” (i.e., an event in which a woman nearly dies), clinically recognized as severe maternal morbidity (5) (SMM). SMM is characterized as a diverse group of life-threatening conditions that arise during labor and delivery and disproportionally affects racial and ethnic minority birthing populations.(6) The Centers for Disease Prevention and Control (CDC) currently monitors national and state-level trends in SMM (7). SMM affects residents in rural communities (8), women with comorbidities (9) and older mothers (8). Black women are much more likely than white women to suffer from these life-threatening complications (10).

The risk of SMM is much higher in the Southern United States (US), particularly in South Carolina (SC) and among Black women (SC) (11, 12). Individual SMM indicators, like eclampsia and pregnancy-related hypertension, have been connected to environmental phenomena such as heatwaves and seasonal air pollution (13, 14). One recent study showed that the likelihood of residing in a high-risk SMM cluster was much higher for women who experienced the most number of hot and very hot days during their pregnancy (15). But the impact of cold and heat-related temperature exposures on elevated SMM risk has not yet been wholly evaluated. The objective of this retrospective birth cohort study was to characterize the association between cold and heat-related temperature extremes and differential SMM risks across critical windows of pregnancy in a US-based Southern cohort.

## Subjects and Methods

### Retrospective cohort description

A retrospective cohort design was used to examine the differential risk of SMM in the presence of extreme ambient temperature by linking hospital-delivery discharge, birth, and maternal mortality records for all deliveries in SC from 1999 to 2018. Our sample was restricted to all singleton births between 20 and 42 weeks of gestation. Institutional Review Board approval was obtained (IRB# 24297).

### Severe Maternal Morbidity

A published algorithm (7) was used to identify daily counts of SMM, a composite metric that includes 21 indicators of severe maternal complications (Table S1). The recent rise in SMM rates nationally are due to the increasing prevalence of blood transfusions (5). One important limitation is that hospital administrative records do not include the number of units of blood transferred and may result in artificially high SMM rates. Therefore, we examined SMM without blood transfusion as the primary outcome of interest. All administrative data using ICD-10 codes from October 2015 to 2018 were back coded to ICD-9 coding equivalents using published general equivalence mapping (16). The CDC’s definition of SMM has been demonstrated to perform as well as chart reviews, the current gold standard for validation (17).

**Table 1.**
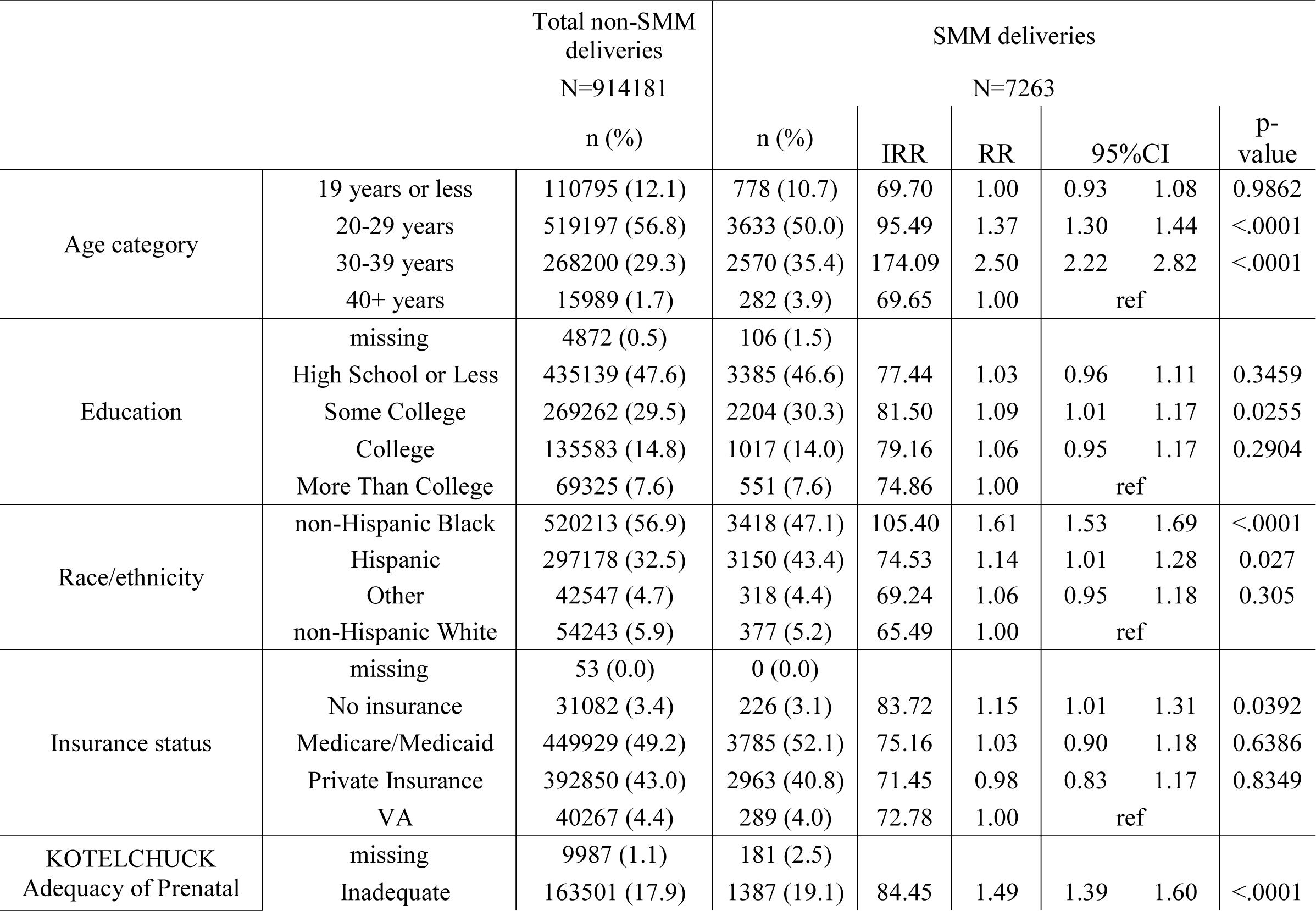
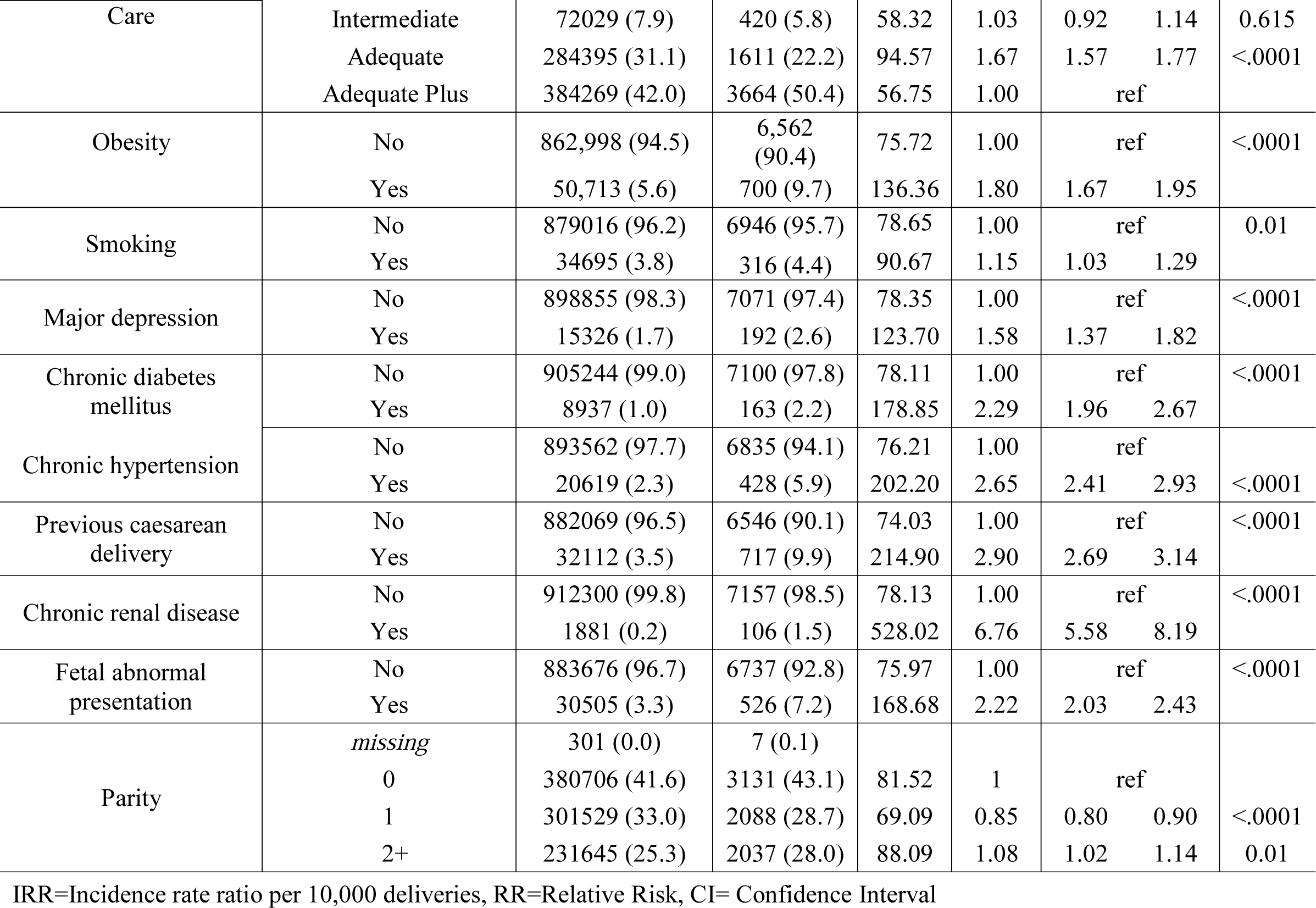
Sample characteristics and incidence of SMM deliveries, South Carolina 1999-2017.

### Critical time windows

To define critical windows of pregnancy, the estimated day of conception was calculated by subtracting the hospital admission date from gestational age. We calculated five gestational windows of exposure: 1) *preconception* (12 weeks before conception) (Pre); 2) *trimester 1* (1-13 gestation weeks) (T1); 3) *trimester 2* (14-26 gestation weeks) (T2); and 4) *trimester 3* (27+ gestation weeks) (T3). Fitting regression models stratified by gestational period is similar to a pregnancy-at-risk approach (18, 19) and relative risk (RR) estimates can be conceptualized as the risk of delivery for a given gestational period. The count of days for each temperature threshold within each critical period of pregnancy (e.g., 12 days of very cold (<5^th^ percentile) occurred in T1) was included rather than the average temperature to avoid masking the variation of daily temperature exposure (20) and reflect local acclimation experienced across the critical periods.

### Temperature exposure assessment

Daily data on the county-level mean (Tavg) (i.e., magnitude of temperature experienced on average during the day) temperature was obtained from NOAA’s nClimGrid (21).

Temperature was examined across the entire year rather than restrict to the cold period (Oct-Feb) or hot period (May-Sept). Adjusting for the season as a covariate may result in potential collinearity between seasons and temperature; therefore, the primary analysis did not adjust for the season to better examine the role of ambient temperature (22). Ambient temperature rather than humidity, heat index, or other weather variables was used because the majority of environmental health studies have shown that temperature is the predominant meteorological factor associated with adverse health outcomes (23).

Exposure was defined as the count of days for the following percentile-based indices of temperature extremes: *very cold* (<5th percentile), *cold* (5th and 10th percentile), and *hot* (90th and 95th percentile), and *very hot* (above 95th percentile) (22, 24) for each critical time window.

Because each exposure threshold index had a large number of zeros for days of cold or hot extremes during each trimester (Table S2), we examined three category levels of exposure for each temperature extreme metric separately to reduce the potential for bias: (1)’no exposure’ (e.g., 0 days in which temperatures reached an extreme); (2) ‘low exposure’ (defined as number of total days <90th percentile for heat extremes or number of total days <10^th^ percentile for cold extremes) and (3) ‘high exposure’ (defined as total number of days >= 90th percentile for heat extremes or number of total days <= 10th percentile for cold extremes, respectively, based on county of maternal residence.

**Table 2.**
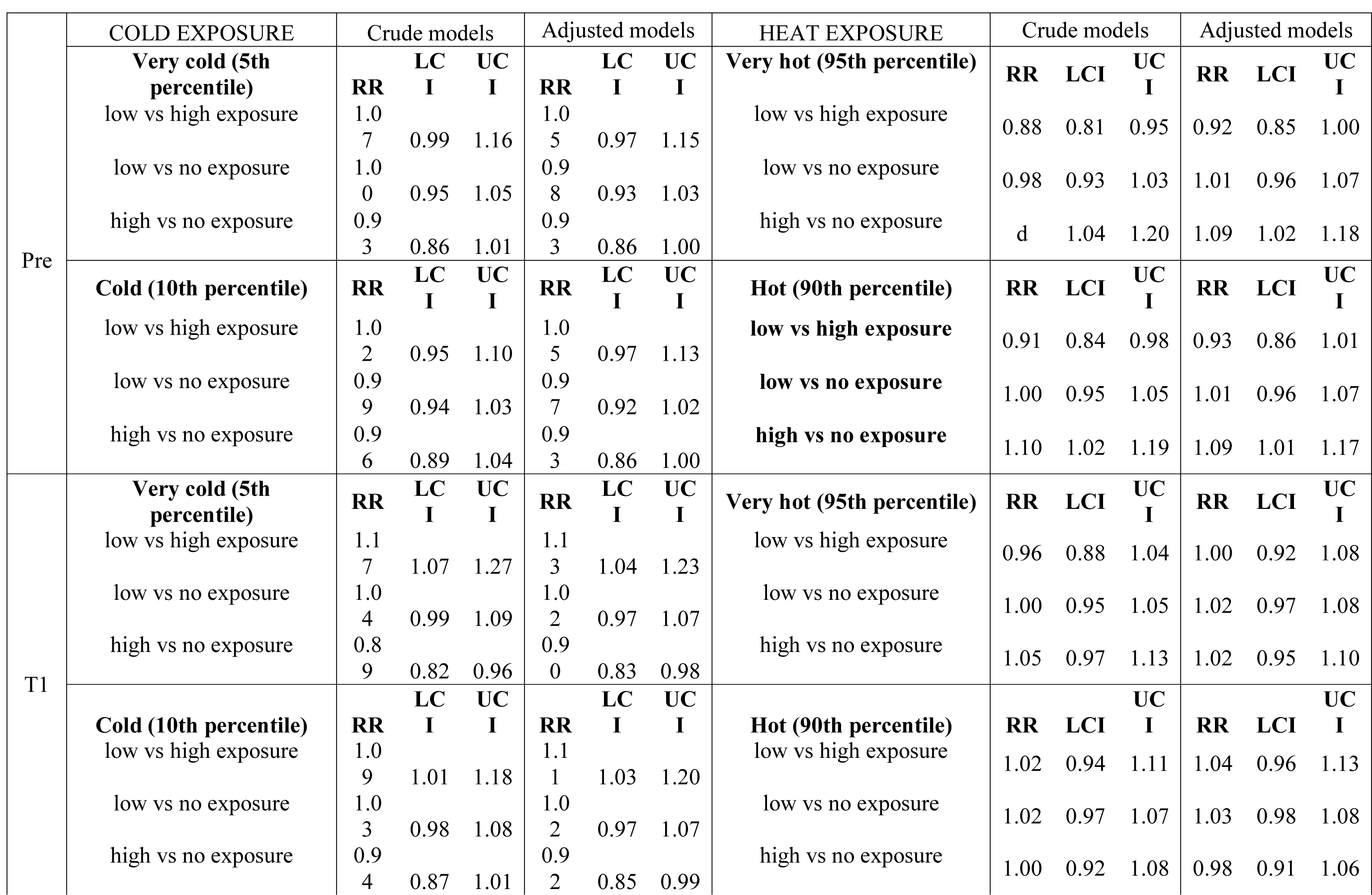
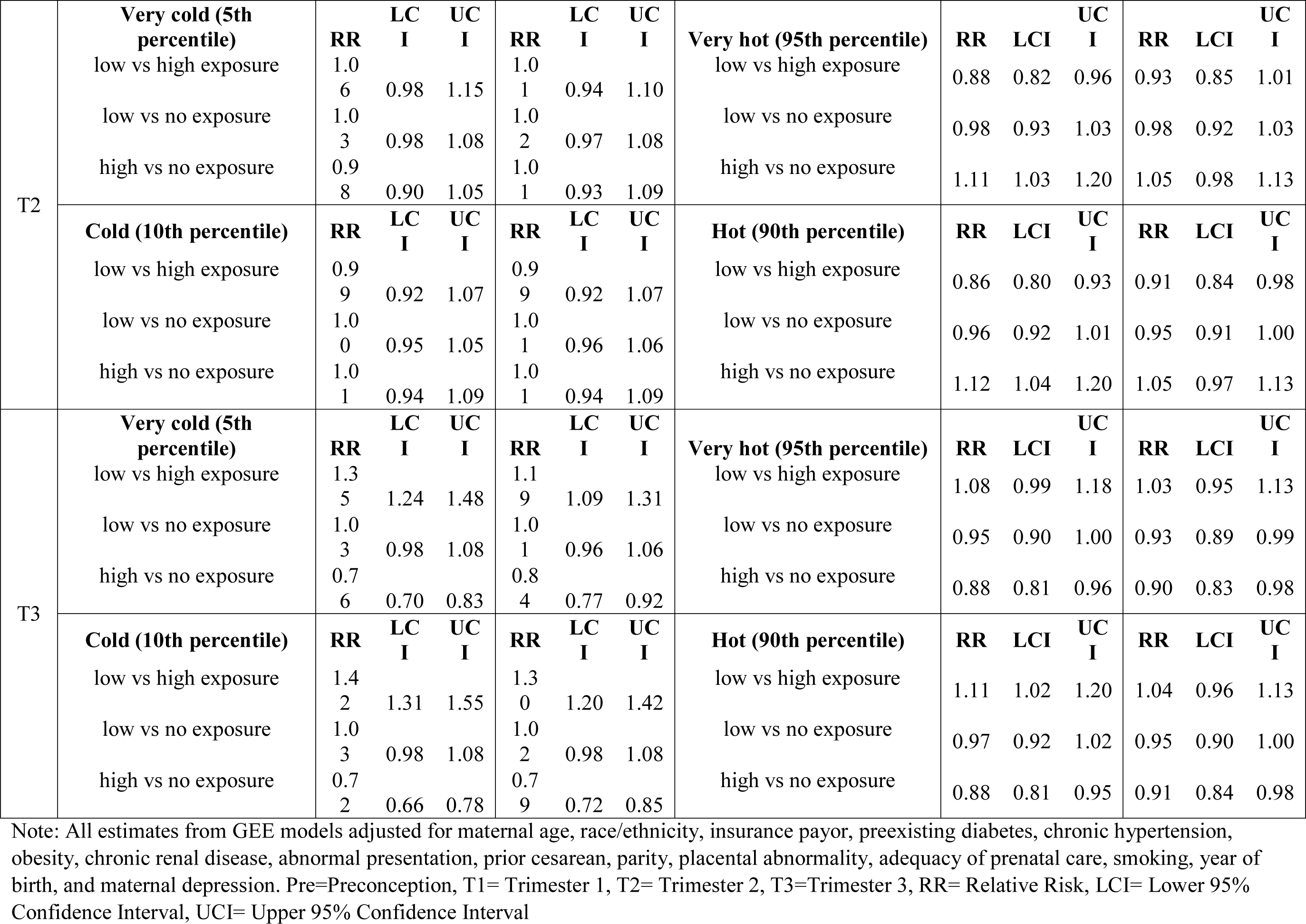
Crude and adjusted relative risk (RR) for cold and hot temperature extremes across critical periods of pregnancy, South Carolina 1999-2017.

To assess heatwave exposure, we included the excess heat factor (EHF) index to examine thresholds of vulnerability to low, moderate, and high-intensity heat waves (25). The EHF is a new metric that takes into consideration the county-level local climate and 30-day acclimatization. Heat wave conditions were then categorized as: (1) *no heatwave*: daily EHF_sev_ ≤ 0; (2) *low intensity*: daily EHF_sev_ ≤ 0 and < 1; (3) *moderate severity*: daily EHF_sev_ ≥ 1 and < 2; and (4) *high severity*: daily EHF_sev_ ≥ 2 (25, 26) for each critical period of pregnancy.

### Covariates

The following potential confounders were abstracted from hospital discharge records and included in the analysis: maternal age (<19 years, 20-29 years, 30-39 years, 40+ years), race/ethnicity (non-Hispanic Black, non-Hispanic White, Hispanic, Other), insurance (Medicare/Medicaid, Private Insurance, Self-pay, Other), education status (high school or less, some college, college, more than college, college), chronic hypertension (ICD9: 401.0,401.1 or 401.9), obesity (ICD9: 278), cesarean delivery (ICD9: 654.20), abnormal presentation (ICD9: 652.2), Type 2 diabetes mellitus (ICD9: 250), parity (none, one previous live birth, two or more), year of birth, placental disorder (presence of placenta previa, placenta accreta, placenta increta or placenta percreta), chronic renal disease (585.9), and Kotelchuck index, a proxy for Adequacy of Prenatal Care Utilization (27–32). BMI was only available starting in 2004 and was not included in the analysis. Lastly, we used 2013 Rural-Urban Continuum codes to define maternal residence in either urban or rural counties (33).

### Other socio-environmental environmental stressors: Air quality, Poverty, and Structural Racism

Poor AQ has also been cited as a potential confounder in temperature-maternal health relationships (34, 35). Daily AQ was obtained from the US Environmental Protection Agency (EPA) Fused Air Quality Surface using Downscaling data and was calculated as days of Ozone (ppm) 0.071-0.085 deemed unhealthy for sensitive groups and included as a confounder (36).

Residential segregation (a proxy for structural racism) and poverty have emerged as two significant drivers that may in part explain differential vulnerability to environmental hazards, particularly for racially and ethnically diverse pregnant populations (37). County-level data on racialized economic segregation (e.g., racial composition, household income) was extracted from the Decennial Census Bureau and matched with deliveries for the 2000 (deliveries between 1999 and 2009) and 2010 estimates (deliveries between 2010 and 2017). The Index of the Concentration of Extremes (ICE) to compare temperature-SMM risks between tertiles of racially and economically segregated communities (ICE_race&income_) (i.e., Q1: low-income, majority Black compared to Q3: high-income, majority-white) (38).

### Statistical Analysis

Generalized estimating equations (GEE) using a modified Poisson approach was used for our correlated SMM data to account for clustering among women with multiple pregnancies nested within counties (39, 40). We used generalized estimating equations (GEE) to adjust for multiple pregnancies for each woman (41) and a Poisson distribution with link function was used to estimate the relative risk (RR) of SMM across each exposure window and temperature extreme (42). The crude RR and adjusted RR with 95% confidence intervals were calculated to estimate the strength of the association between temperature extremes and SMM. The following covariates were adjusted for in all final models: maternal age, race/ethnicity, insurance payor, preexisting diabetes, chronic hypertension, obesity, chronic renal disease, abnormal presentation, prior cesarean, parity, placental abnormality, adequacy of prenatal care, smoking, year of birth, and maternal depression.

### Sensitivity analysis

We conducted the following sensitivity analysis: 1) adjusted for ozone exposure over each critical period in full temperature models; 2) examined heat waves and their associated intensity (i.e., low, moderate, and high severity) in the warm season across each critical period; and 3) examined rural vs urban, maternal age (<35 years, ≥ 35 years), preterm birth (live birth occurring at < 37 weeks of gestation as noted on the birth certificate), and ICE_race&income_ as effect modifiers using as an interaction term with Bonferroni adjustments.

### Excess SMM cases due to temperature extremes

The attribution of climate-related change in temperature contributing to excess SMM can be estimated by examining the attributable risk of temperature in the exposed SMM cases compared to unexposed SMM cases, a similar approach used by climate scientists to attribute climate change to extreme weather events using counterfactual scenarios (43). The unadjusted excess fraction (EF), a proxy to attributable risk in the absence of potential completion time of sufficient causes, was calculated to estimate the proportion of excess incident SMM cases due to exposure to hot or cold ambient extremes (44). Finally, the population attributable risk (PAR) was calculated to estate the proportion of SMM risk in the total population associated with exposure to either cold or hot temperatures (44). Proc stdrate in SAS 9.4 using the Mantel-Haenszel Estimation method to generate the EF and PAR and associated 95% confidence intervals (45).

## Results

### Descriptive characteristics

Table 1 shows maternal demographic characteristics. There were 921,444 delivery hospitalizations and a total of 7,263 SMM deliveries without blood transfusion for the study period. SMM deliveries were more prevalent among persons who were older (30-39 years), non-Hispanic Black or Hispanic, lacked insurance, had placental issues, experienced inadequate or adequate plus prenatal care, had some college education, and among smokers. SMM deliveries were also more common associated with previous cesarean delivery, obesity, depression, chronic hypertension, renal disease or diabetes, and an infant with an abnormal presentation at delivery. Figure 1 demonstrates that the incidence rate of SMM in SC has been on the rise since 1999.

**Figure 1.**
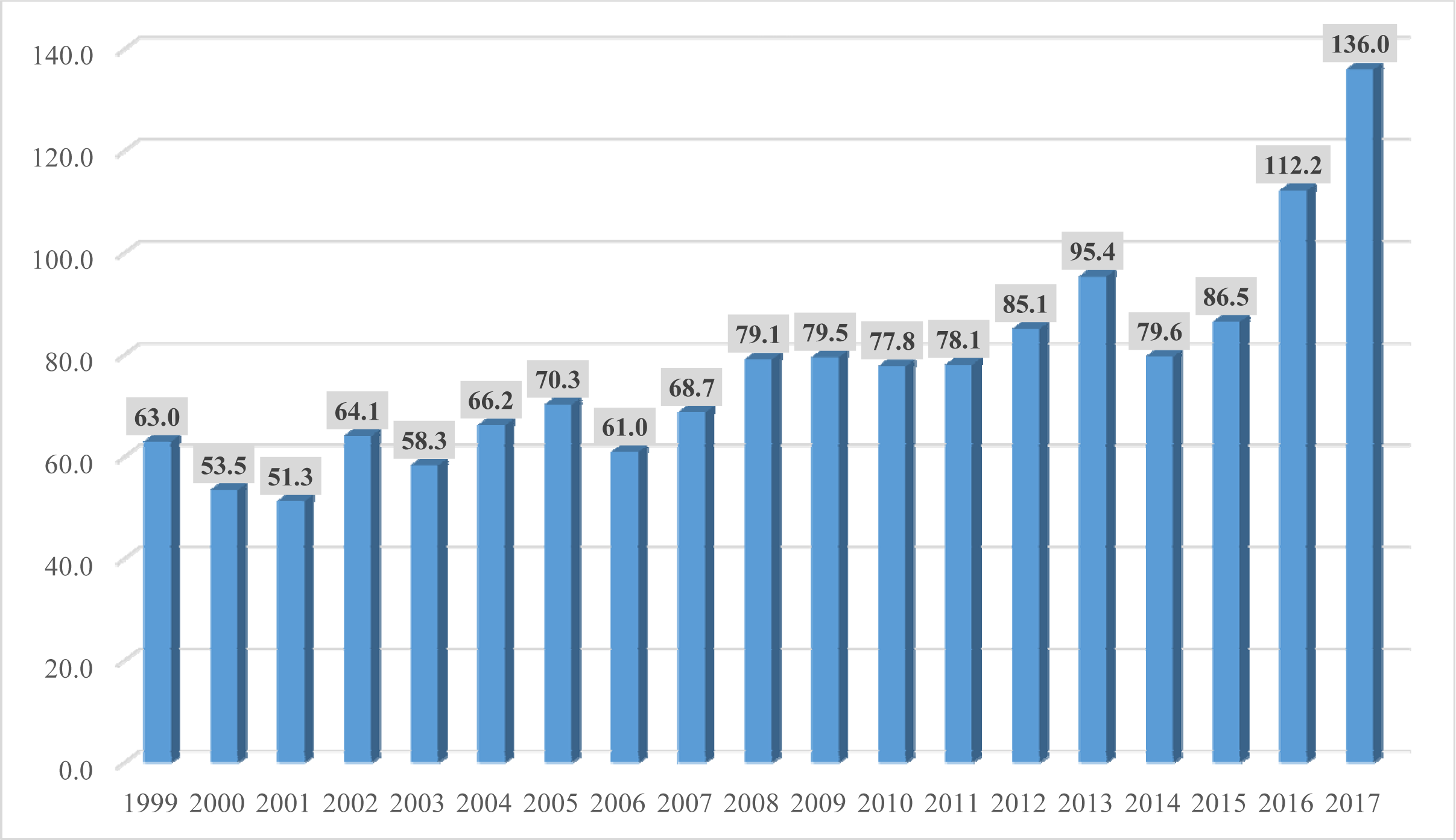
Annual incidence rate ratio for severe maternal morbidity per 10,000 delivery hospitalizations, South Carolina 1999-2017.

### Prenatal exposure to cold ambient extremes

A crude association between cold and very cold temperature exposure and elevated SMM risk was observed during the T1 and T3 (Table 2). Adjusted models demonstrated that exposure to low compared to a high number of cold and very cold temperatures during T1 and T3 was associated with an 11% to 30% increase, respectively, in SMM risks during delivery (Figure 2). There was no difference in SMM risks for exposure to either cold or very cold temperatures detected for T1, suggesting that even a few days of exposure to colder than average temperatures was a risk factor of SMM.

**Figure 2.**
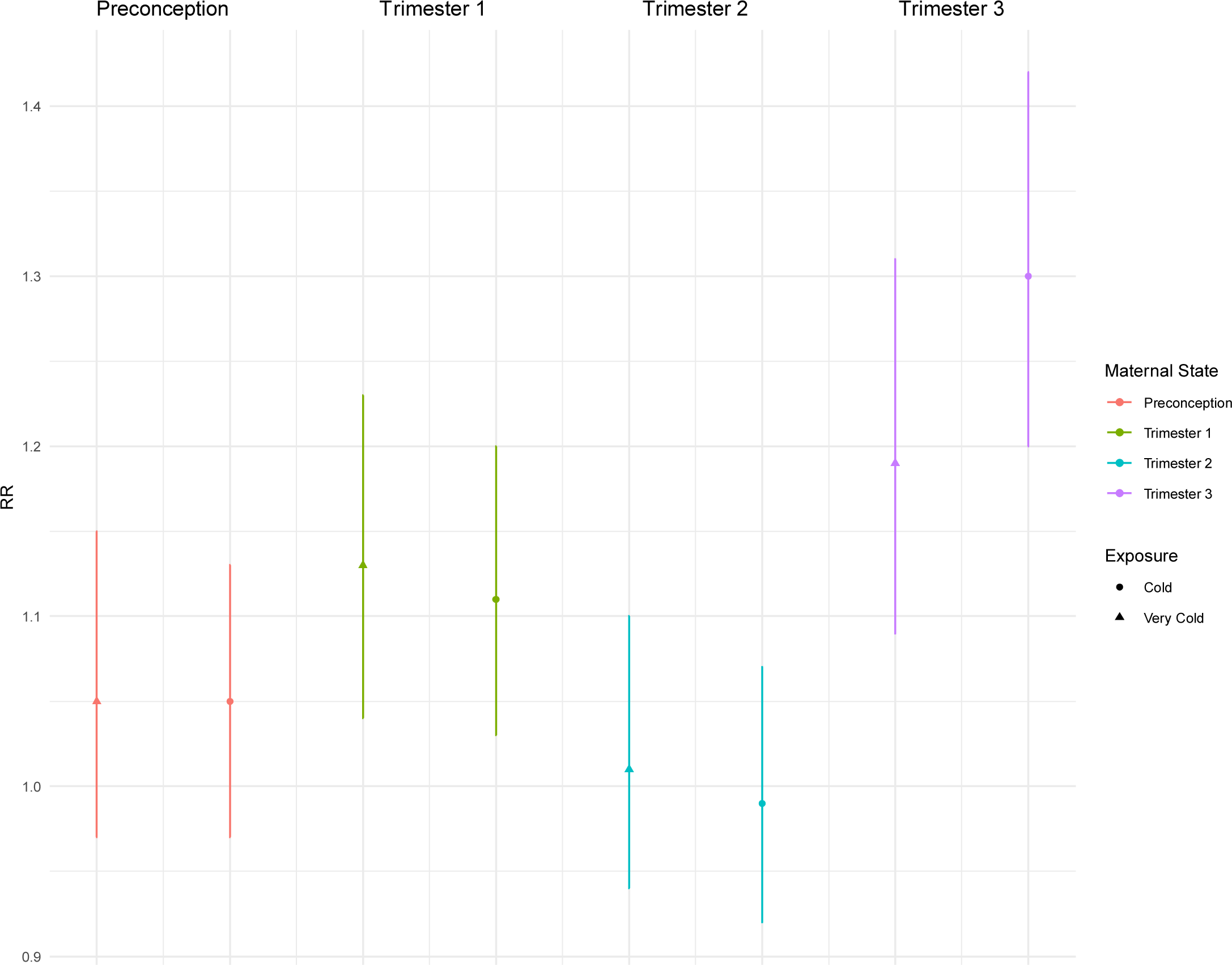
Adjusted relative risk with 95% confidence intervals showing the association between maternal exposure to low compared to a high number of cold and very cold days for each critical period.

### Prenatal exposure to hot ambient extremes

Crude estimates revealed increased SMM risks for pregnant persons exposed to very hot and hot temperatures during the Pre period and the T2 and T3 (Table 2). After adjustment for important sociodemographic and maternal/obstetric covariates, women exposed to high ambient temperatures (>=90th percentile) during preconception were 9% more likely to experience an SMM event compared to women not exposed (Figure 3). After restricting to the summer season, results showed a two-fold risk in SMM following maternal exposure to hot and very hot temperatures in T2 compared to no exposure to hot temperatures (Table S3).

**Figure 3.**
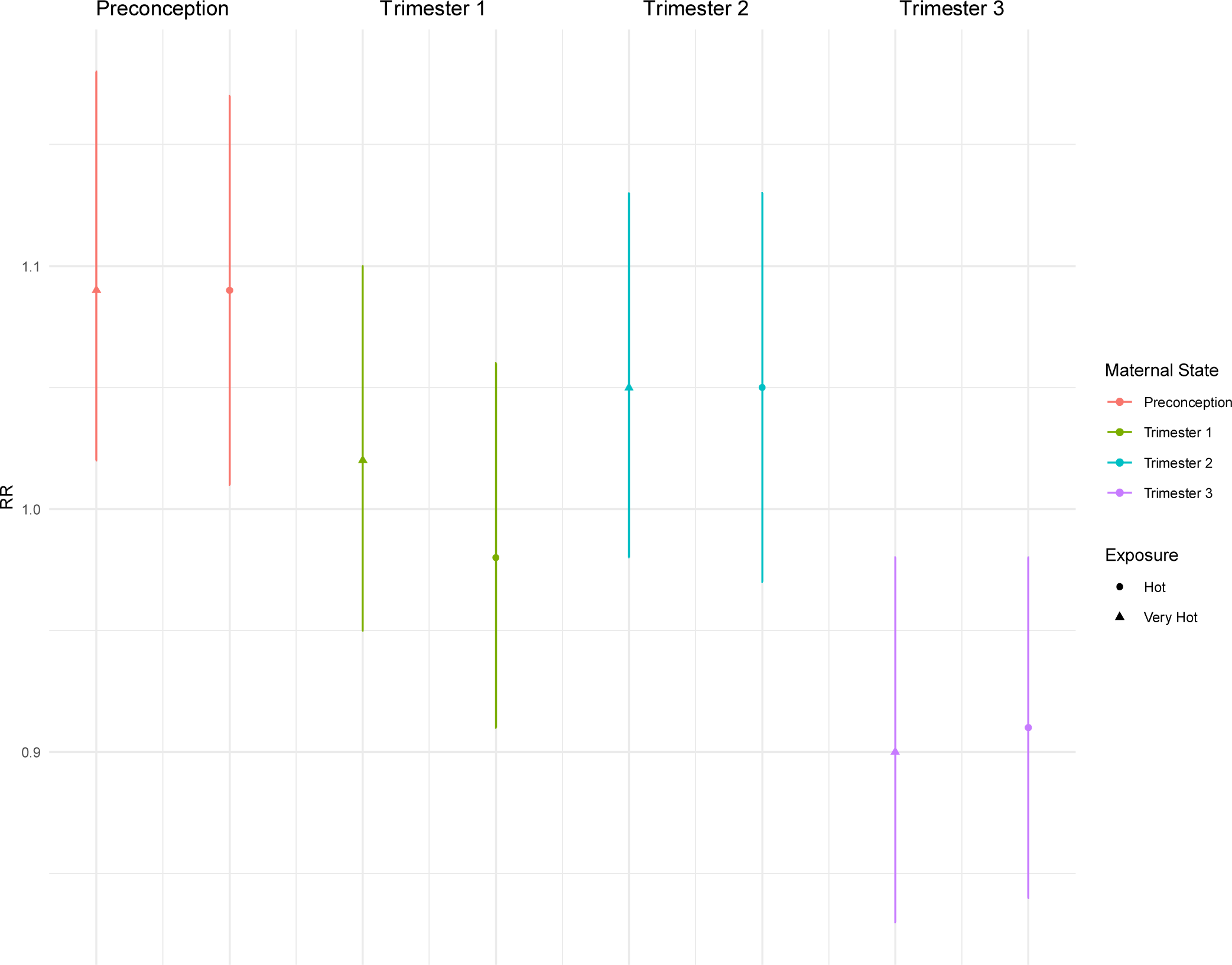
Adjusted relative risk with 95% confidence intervals showing the association between a high number of hot and very hot days compared to no exposure for each critical period.

**Table 3.**
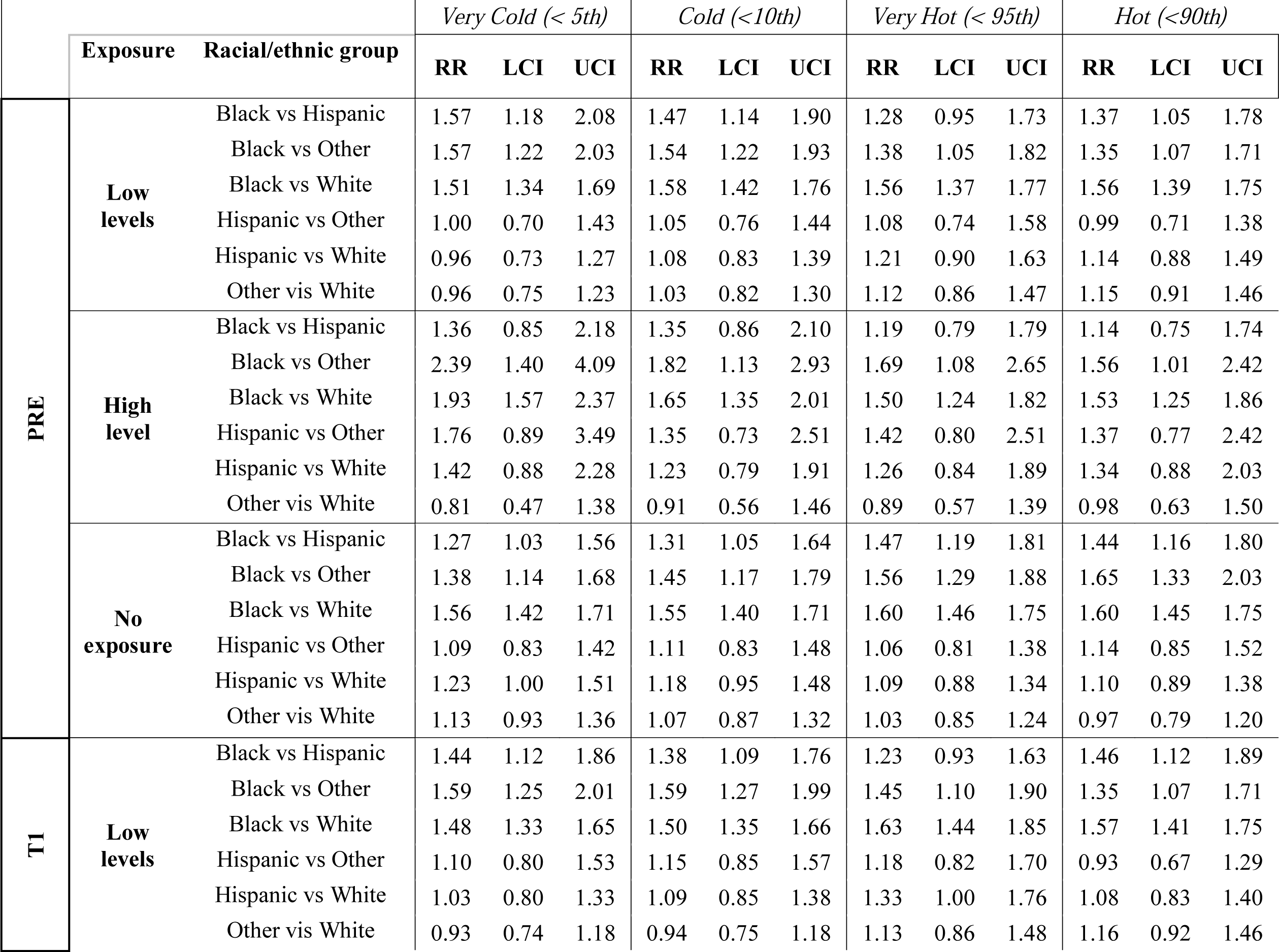
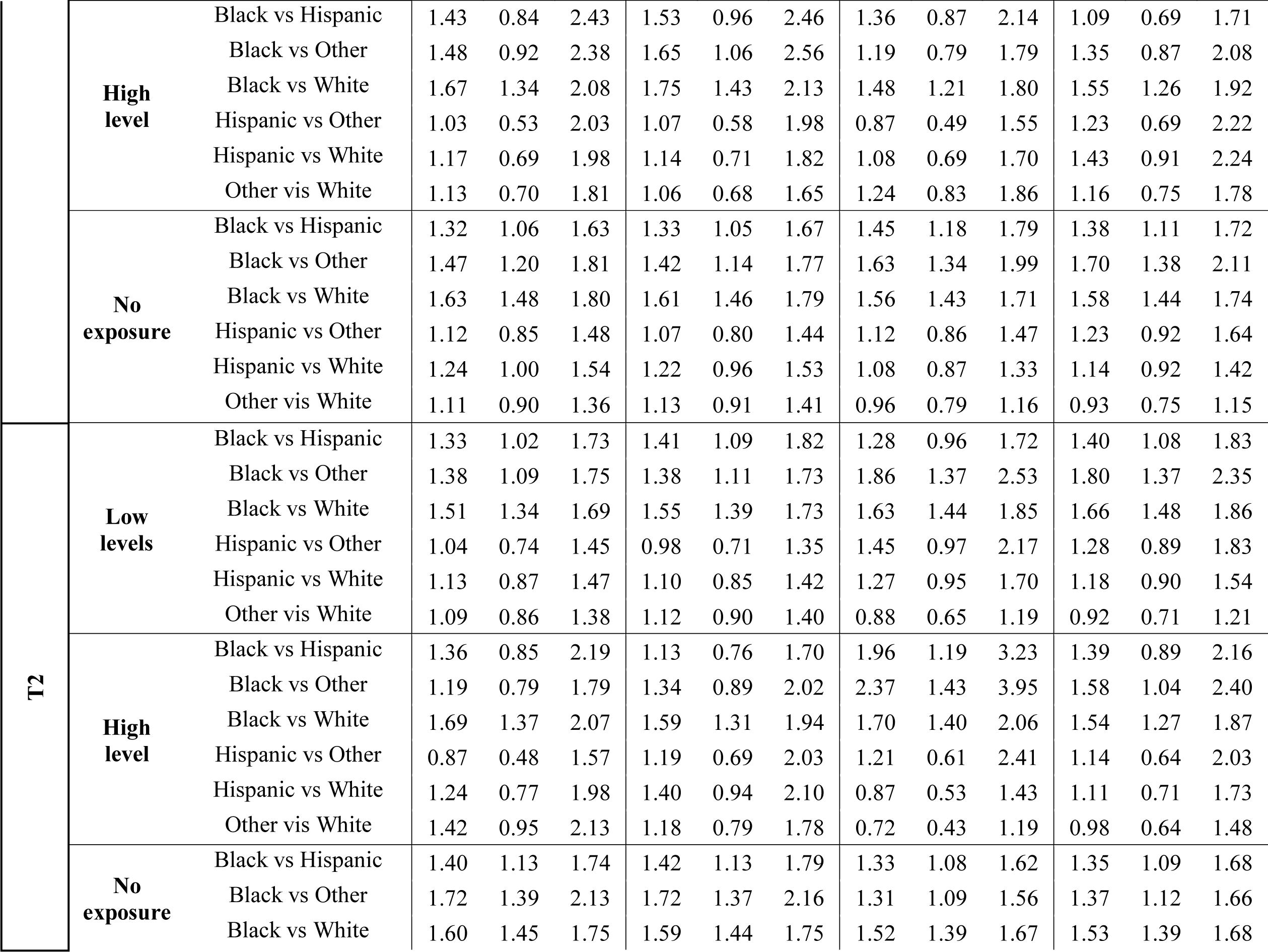
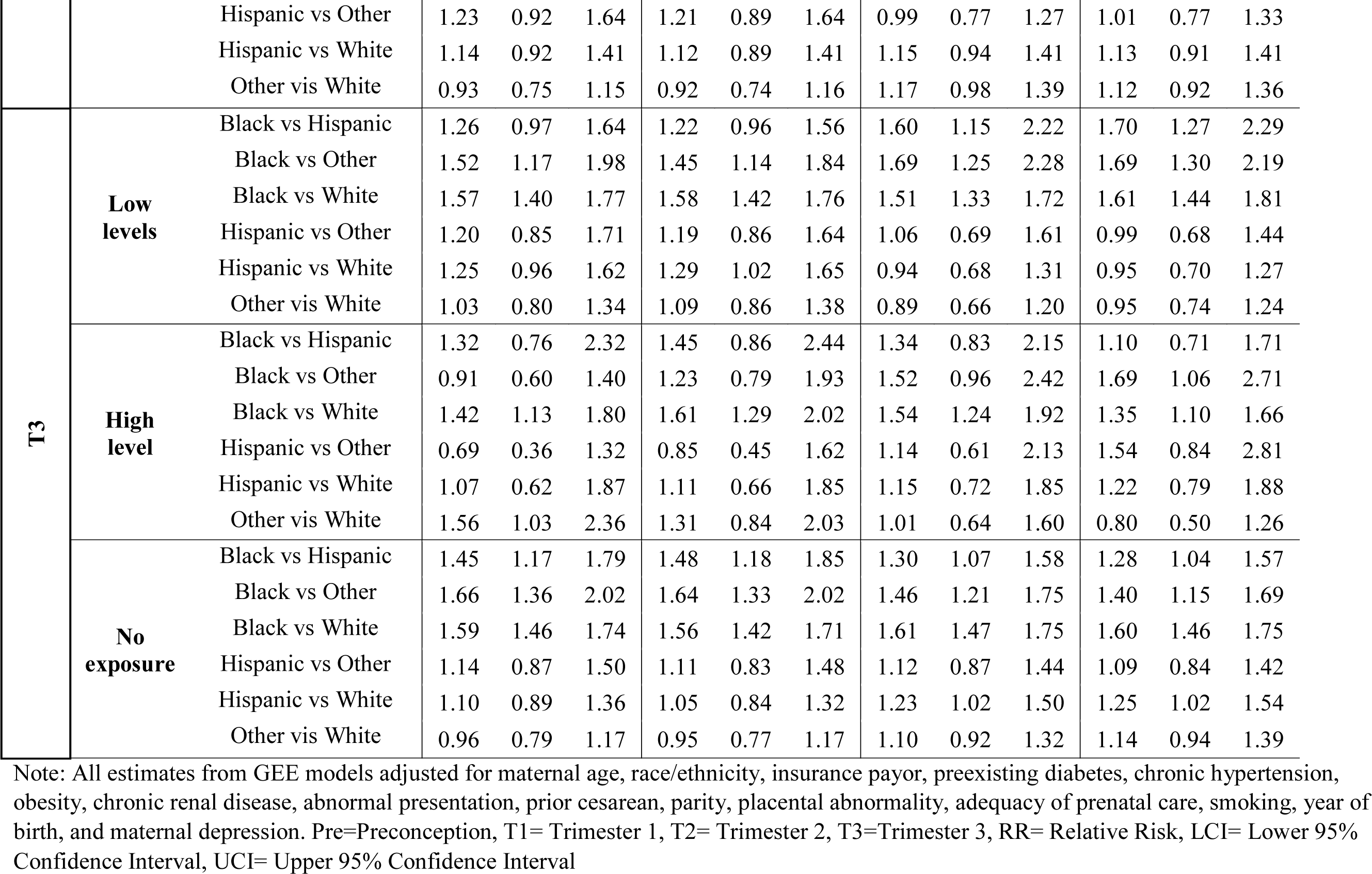
Racial and ethnic differences in SMM risks across levels of cold and hot temperature exposure for critical periods of pregnancy.

|  | Exposure | Racial/ethnic group | Very Cold (< 5th) |  |  | Cold (<10th) |  |  | Very Hot (< 95th) |  |  | Hot (<90th) |  |  |
| --- | --- | --- | --- | --- | --- | --- | --- | --- | --- | --- | --- | --- | --- | --- |
|  |  |  | RR | LCI | UCI | RR | LCI | UCI | RR | LCI | UCI | RR | LCI | UCI |
| PRE | Low levels | Black vs Hispanic | 1.57 | 1.18 | 2.08 | 1.47 | 1.14 | 1.90 | 1.28 | 0.95 | 1.73 | 1.37 | 1.05 | 1.78 |
|  |  | Black vs Other | 1.57 | 1.22 | 2.03 | 1.54 | 1.22 | 1.93 | 1.38 | 1.05 | 1.82 | 1.35 | 1.07 | 1.71 |
|  |  | Black vs White | 1.51 | 1.34 | 1.69 | 1.58 | 1.42 | 1.76 | 1.56 | 1.37 | 1.77 | 1.56 | 1.39 | 1.75 |
|  |  | Hispanic vs Other | 1.00 | 0.70 | 1.43 | 1.05 | 0.76 | 1.44 | 1.08 | 0.74 | 1.58 | 0.99 | 0.71 | 1.38 |
|  |  | Hispanic vs White | 0.96 | 0.73 | 1.27 | 1.08 | 0.83 | 1.39 | 1.21 | 0.90 | 1.63 | 1.14 | 0.88 | 1.49 |
|  |  | Other vis White | 0.96 | 0.75 | 1.23 | 1.03 | 0.82 | 1.30 | 1.12 | 0.86 | 1.47 | 1.15 | 0.91 | 1.46 |
|  | High level | Black vs Hispanic | 1.36 | 0.85 | 2.18 | 1.35 | 0.86 | 2.10 | 1.19 | 0.79 | 1.79 | 1.14 | 0.75 | 1.74 |
|  |  | Black vs Other | 2.39 | 1.40 | 4.09 | 1.82 | 1.13 | 2.93 | 1.69 | 1.08 | 2.65 | 1.56 | 1.01 | 2.42 |
|  |  | Black vs White | 1.93 | 1.57 | 2.37 | 1.65 | 1.35 | 2.01 | 1.50 | 1.24 | 1.82 | 1.53 | 1.25 | 1.86 |
|  |  | Hispanic vs Other | 1.76 | 0.89 | 3.49 | 1.35 | 0.73 | 2.51 | 1.42 | 0.80 | 2.51 | 1.37 | 0.77 | 2.42 |
|  |  | Hispanic vs White | 1.42 | 0.88 | 2.28 | 1.23 | 0.79 | 1.91 | 1.26 | 0.84 | 1.89 | 1.34 | 0.88 | 2.03 |
|  |  | Other vis White | 0.81 | 0.47 | 1.38 | 0.91 | 0.56 | 1.46 | 0.89 | 0.57 | 1.39 | 0.98 | 0.63 | 1.50 |
|  | No exposure | Black vs Hispanic | 1.27 | 1.03 | 1.56 | 1.31 | 1.05 | 1.64 | 1.47 | 1.19 | 1.81 | 1.44 | 1.16 | 1.80 |
|  |  | Black vs Other | 1.38 | 1.14 | 1.68 | 1.45 | 1.17 | 1.79 | 1.56 | 1.29 | 1.88 | 1.65 | 1.33 | 2.03 |
|  |  | Black vs White | 1.56 | 1.42 | 1.71 | 1.55 | 1.40 | 1.71 | 1.60 | 1.46 | 1.75 | 1.60 | 1.45 | 1.75 |
|  |  | Hispanic vs Other | 1.09 | 0.83 | 1.42 | 1.11 | 0.83 | 1.48 | 1.06 | 0.81 | 1.38 | 1.14 | 0.85 | 1.52 |
|  |  | Hispanic vs White | 1.23 | 1.00 | 1.51 | 1.18 | 0.95 | 1.48 | 1.09 | 0.88 | 1.34 | 1.10 | 0.89 | 1.38 |
|  |  | Other vis White | 1.13 | 0.93 | 1.36 | 1.07 | 0.87 | 1.32 | 1.03 | 0.85 | 1.24 | 0.97 | 0.79 | 1.20 |
| T1 | Low levels | Black vs Hispanic | 1.44 | 1.12 | 1.86 | 1.38 | 1.09 | 1.76 | 1.23 | 0.93 | 1.63 | 1.46 | 1.12 | 1.89 |
|  |  | Black vs Other | 1.59 | 1.25 | 2.01 | 1.59 | 1.27 | 1.99 | 1.45 | 1.10 | 1.90 | 1.35 | 1.07 | 1.71 |
|  |  | Black vs White | 1.48 | 1.33 | 1.65 | 1.50 | 1.35 | 1.66 | 1.63 | 1.44 | 1.85 | 1.57 | 1.41 | 1.75 |
|  |  | Hispanic vs Other | 1.10 | 0.80 | 1.53 | 1.15 | 0.85 | 1.57 | 1.18 | 0.82 | 1.70 | 0.93 | 0.67 | 1.29 |
|  |  | Hispanic vs White | 1.03 | 0.80 | 1.33 | 1.09 | 0.85 | 1.38 | 1.33 | 1.00 | 1.76 | 1.08 | 0.83 | 1.40 |
|  |  | Other vis White | 0.93 | 0.74 | 1.18 | 0.94 | 0.75 | 1.18 | 1.13 | 0.86 | 1.48 | 1.16 | 0.92 | 1.46 |

|  |  |  |  |  |  |  |  |  |  |  |  |  |  |  |  |
| --- | --- | --- | --- | --- | --- | --- | --- | --- | --- | --- | --- | --- | --- | --- | --- |
|  |  | **High level** | Black vs Hispanic | 1.43 | 0.84 | 2.43 | 1.53 | 0.96 | 2.46 | 1.36 | 0.87 | 2.14 | 1.09 | 0.69 | 1.71 |
|  |  |  | Black vs Other | 1.48 | 0.92 | 2.38 | 1.65 | 1.06 | 2.56 | 1.19 | 0.79 | 1.79 | 1.35 | 0.87 | 2.08 |
|  |  |  | Black vs White | 1.67 | 1.34 | 2.08 | 1.75 | 1.43 | 2.13 | 1.48 | 1.21 | 1.80 | 1.55 | 1.26 | 1.92 |
|  |  |  | Hispanic vs Other | 1.03 | 0.53 | 2.03 | 1.07 | 0.58 | 1.98 | 0.87 | 0.49 | 1.55 | 1.23 | 0.69 | 2.22 |
|  |  |  | Hispanic vs White | 1.17 | 0.69 | 1.98 | 1.14 | 0.71 | 1.82 | 1.08 | 0.69 | 1.70 | 1.43 | 0.91 | 2.24 |
|  |  |  | Other vis White | 1.13 | 0.70 | 1.81 | 1.06 | 0.68 | 1.65 | 1.24 | 0.83 | 1.86 | 1.16 | 0.75 | 1.78 |
|  |  | **No exposure** | Black vs Hispanic | 1.32 | 1.06 | 1.63 | 1.33 | 1.05 | 1.67 | 1.45 | 1.18 | 1.79 | 1.38 | 1.11 | 1.72 |
|  |  |  | Black vs Other | 1.47 | 1.20 | 1.81 | 1.42 | 1.14 | 1.77 | 1.63 | 1.34 | 1.99 | 1.70 | 1.38 | 2.11 |
|  |  |  | Black vs White | 1.63 | 1.48 | 1.80 | 1.61 | 1.46 | 1.79 | 1.56 | 1.43 | 1.71 | 1.58 | 1.44 | 1.74 |
|  |  |  | Hispanic vs Other | 1.12 | 0.85 | 1.48 | 1.07 | 0.80 | 1.44 | 1.12 | 0.86 | 1.47 | 1.23 | 0.92 | 1.64 |
|  |  |  | Hispanic vs White | 1.24 | 1.00 | 1.54 | 1.22 | 0.96 | 1.53 | 1.08 | 0.87 | 1.33 | 1.14 | 0.92 | 1.42 |
|  |  |  | Other vis White | 1.11 | 0.90 | 1.36 | 1.13 | 0.91 | 1.41 | 0.96 | 0.79 | 1.16 | 0.93 | 0.75 | 1.15 |
| **T2** |  | **Low levels** | Black vs Hispanic | 1.33 | 1.02 | 1.73 | 1.41 | 1.09 | 1.82 | 1.28 | 0.96 | 1.72 | 1.40 | 1.08 | 1.83 |
|  |  |  | Black vs Other | 1.38 | 1.09 | 1.75 | 1.38 | 1.11 | 1.73 | 1.86 | 1.37 | 2.53 | 1.80 | 1.37 | 2.35 |
|  |  |  | Black vs White | 1.51 | 1.34 | 1.69 | 1.55 | 1.39 | 1.73 | 1.63 | 1.44 | 1.85 | 1.66 | 1.48 | 1.86 |
|  |  |  | Hispanic vs Other | 1.04 | 0.74 | 1.45 | 0.98 | 0.71 | 1.35 | 1.45 | 0.97 | 2.17 | 1.28 | 0.89 | 1.83 |
|  |  |  | Hispanic vs White | 1.13 | 0.87 | 1.47 | 1.10 | 0.85 | 1.42 | 1.27 | 0.95 | 1.70 | 1.18 | 0.90 | 1.54 |
|  |  |  | Other vis White | 1.09 | 0.86 | 1.38 | 1.12 | 0.90 | 1.40 | 0.88 | 0.65 | 1.19 | 0.92 | 0.71 | 1.21 |
|  |  | **High level** | Black vs Hispanic | 1.36 | 0.85 | 2.19 | 1.13 | 0.76 | 1.70 | 1.96 | 1.19 | 3.23 | 1.39 | 0.89 | 2.16 |
|  |  |  | Black vs Other | 1.19 | 0.79 | 1.79 | 1.34 | 0.89 | 2.02 | 2.37 | 1.43 | 3.95 | 1.58 | 1.04 | 2.40 |
|  |  |  | Black vs White | 1.69 | 1.37 | 2.07 | 1.59 | 1.31 | 1.94 | 1.70 | 1.40 | 2.06 | 1.54 | 1.27 | 1.87 |
|  |  |  | Hispanic vs Other | 0.87 | 0.48 | 1.57 | 1.19 | 0.69 | 2.03 | 1.21 | 0.61 | 2.41 | 1.14 | 0.64 | 2.03 |
|  |  |  | Hispanic vs White | 1.24 | 0.77 | 1.98 | 1.40 | 0.94 | 2.10 | 0.87 | 0.53 | 1.43 | 1.11 | 0.71 | 1.73 |
|  |  |  | Other vis White | 1.42 | 0.95 | 2.13 | 1.18 | 0.79 | 1.78 | 0.72 | 0.43 | 1.19 | 0.98 | 0.64 | 1.48 |
|  |  | **No exposure** | Black vs Hispanic | 1.40 | 1.13 | 1.74 | 1.42 | 1.13 | 1.79 | 1.33 | 1.08 | 1.62 | 1.35 | 1.09 | 1.68 |
|  |  |  | Black vs Other | 1.72 | 1.39 | 2.13 | 1.72 | 1.37 | 2.16 | 1.31 | 1.09 | 1.56 | 1.37 | 1.12 | 1.66 |
|  |  |  | Black vs White | 1.60 | 1.45 | 1.75 | 1.59 | 1.44 | 1.75 | 1.52 | 1.39 | 1.67 | 1.53 | 1.39 | 1.68 |
|  |  | Hispanic vs Other | 1.23 | 0.92 | 1.64 | 1.21 | 0.89 | 1.64 | 0.99 | 0.77 | 1.27 | 1.01 | 0.77 | 1.33 |
|  |  | Hispanic vs White | 1.14 | 0.92 | 1.41 | 1.12 | 0.89 | 1.41 | 1.15 | 0.94 | 1.41 | 1.13 | 0.91 | 1.41 |
|  |  | Other vis White | 0.93 | 0.75 | 1.15 | 0.92 | 0.74 | 1.16 | 1.17 | 0.98 | 1.39 | 1.12 | 0.92 | 1.36 |
| **T3** | **Low levels** | Black vs Hispanic | 1.26 | 0.97 | 1.64 | 1.22 | 0.96 | 1.56 | 1.60 | 1.15 | 2.22 | 1.70 | 1.27 | 2.29 |
|  |  | Black vs Other | 1.52 | 1.17 | 1.98 | 1.45 | 1.14 | 1.84 | 1.69 | 1.25 | 2.28 | 1.69 | 1.30 | 2.19 |
|  |  | Black vs White | 1.57 | 1.40 | 1.77 | 1.58 | 1.42 | 1.76 | 1.51 | 1.33 | 1.72 | 1.61 | 1.44 | 1.81 |
|  |  | Hispanic vs Other | 1.20 | 0.85 | 1.71 | 1.19 | 0.86 | 1.64 | 1.06 | 0.69 | 1.61 | 0.99 | 0.68 | 1.44 |
|  |  | Hispanic vs White | 1.25 | 0.96 | 1.62 | 1.29 | 1.02 | 1.65 | 0.94 | 0.68 | 1.31 | 0.95 | 0.70 | 1.27 |
|  |  | Other vis White | 1.03 | 0.80 | 1.34 | 1.09 | 0.86 | 1.38 | 0.89 | 0.66 | 1.20 | 0.95 | 0.74 | 1.24 |
|  | **High level** | Black vs Hispanic | 1.32 | 0.76 | 2.32 | 1.45 | 0.86 | 2.44 | 1.34 | 0.83 | 2.15 | 1.10 | 0.71 | 1.71 |
|  |  | Black vs Other | 0.91 | 0.60 | 1.40 | 1.23 | 0.79 | 1.93 | 1.52 | 0.96 | 2.42 | 1.69 | 1.06 | 2.71 |
|  |  | Black vs White | 1.42 | 1.13 | 1.80 | 1.61 | 1.29 | 2.02 | 1.54 | 1.24 | 1.92 | 1.35 | 1.10 | 1.66 |
|  |  | Hispanic vs Other | 0.69 | 0.36 | 1.32 | 0.85 | 0.45 | 1.62 | 1.14 | 0.61 | 2.13 | 1.54 | 0.84 | 2.81 |
|  |  | Hispanic vs White | 1.07 | 0.62 | 1.87 | 1.11 | 0.66 | 1.85 | 1.15 | 0.72 | 1.85 | 1.22 | 0.79 | 1.88 |
|  |  | Other vis White | 1.56 | 1.03 | 2.36 | 1.31 | 0.84 | 2.03 | 1.01 | 0.64 | 1.60 | 0.80 | 0.50 | 1.26 |
|  | **No exposure** | Black vs Hispanic | 1.45 | 1.17 | 1.79 | 1.48 | 1.18 | 1.85 | 1.30 | 1.07 | 1.58 | 1.28 | 1.04 | 1.57 |
|  |  | Black vs Other | 1.66 | 1.36 | 2.02 | 1.64 | 1.33 | 2.02 | 1.46 | 1.21 | 1.75 | 1.40 | 1.15 | 1.69 |
|  |  | Black vs White | 1.59 | 1.46 | 1.74 | 1.56 | 1.42 | 1.71 | 1.61 | 1.47 | 1.75 | 1.60 | 1.46 | 1.75 |
|  |  | Hispanic vs Other | 1.14 | 0.87 | 1.50 | 1.11 | 0.83 | 1.48 | 1.12 | 0.87 | 1.44 | 1.09 | 0.84 | 1.42 |
|  |  | Hispanic vs White | 1.10 | 0.89 | 1.36 | 1.05 | 0.84 | 1.32 | 1.23 | 1.02 | 1.50 | 1.25 | 1.02 | 1.54 |
|  |  | Other vis White | 0.96 | 0.79 | 1.17 | 0.95 | 0.77 | 1.17 | 1.10 | 0.92 | 1.32 | 1.14 | 0.94 | 1.39 |

Note: All estimates from GEE models adjusted for maternal age, race/ethnicity, insurance payor, preexisting diabetes, chronic hypertension, obesity, chronic renal disease, abnormal presentation, prior cesarean, parity, placental abnormality, adequacy of prenatal care, smoking, year of birth, and maternal depression. Pre=Preconception, T1= Trimester 1, T2= Trimester 2, T3=Trimester 3, RR= Relative Risk, LCI= Lower 95% Confidence Interval, UCI= Upper 95% Confidence Interval

### Racial and ethnic disparities

In general, we observed a higher risk of SMM for Black compared to white women across all levels of cold and hot temperature exposure for each critical period (Table 3). For example, Black women tended to have the highest SMM risk in response to high levels (RR: 1.70, 95%CI:1:40,2.06) of exposure to very hot temperatures in T2 and higher SMM risk in response to high-level exposure to cold temperatures in T1 (RR:1.75, 95%CI:1.43, 2.13) compared to white women.

### Sensitivity Analysis

#### Warm Season Heat Waves

The influence of heatwaves in the warm season (May to Sept) on SMM risks was examine for each critical period. Women exposed to high or moderate intensity heat waves in T2 were 19% more likely to experience SMM (Table 4). Even exposure to low-intensity heat waves during T2 was associated with a 7% increase of a SMM delivery. When only looking at the summer season (Jun-Aug), the risk of a SMM event was even higher following exposure to high and moderate intensity heat waves (RR_high_=1.45, 95%CI: 1.31, 1.60, RR_moderate_=1.39, 95%CI:1.32,1.47).

**Table 4.**
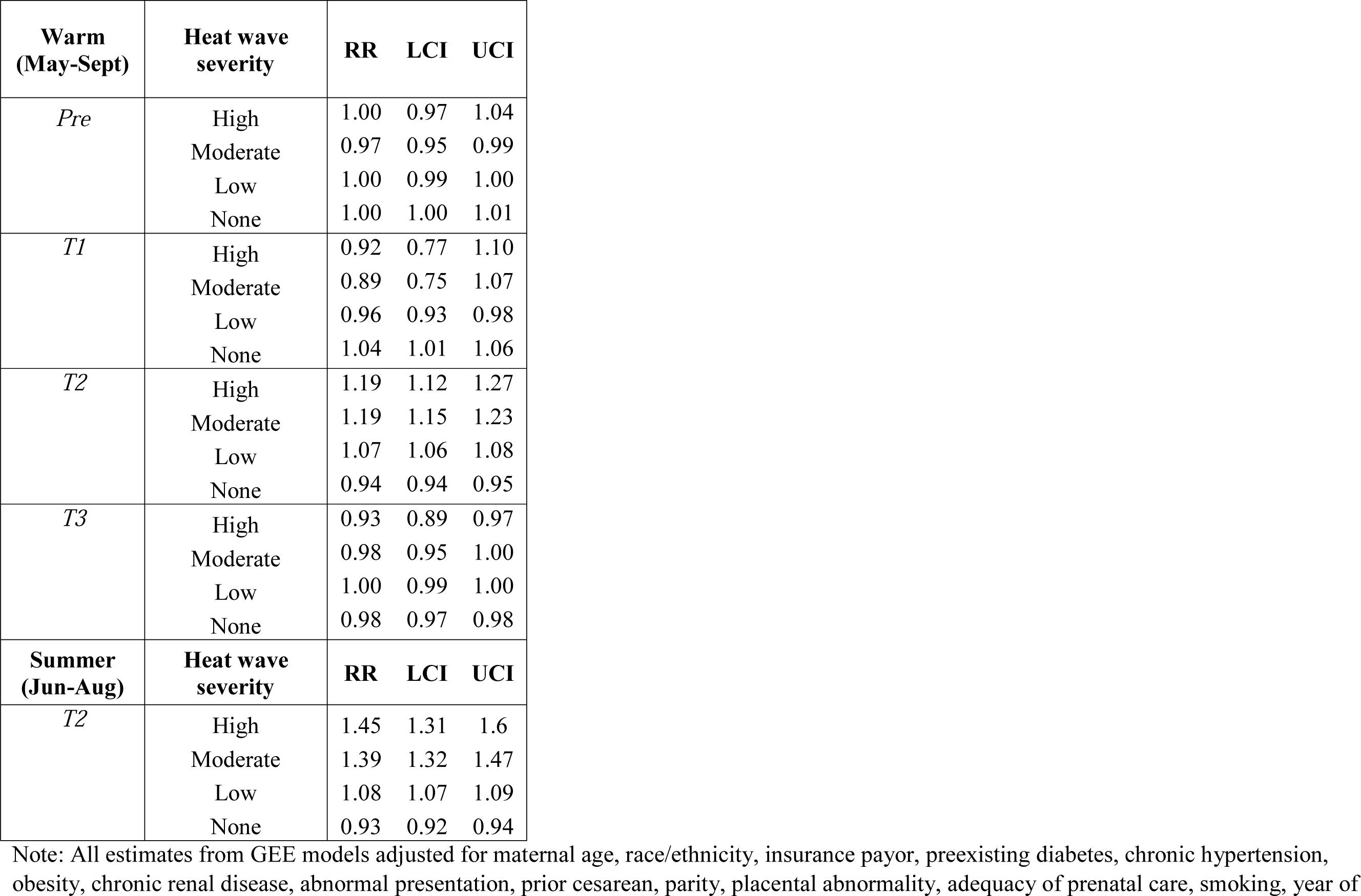
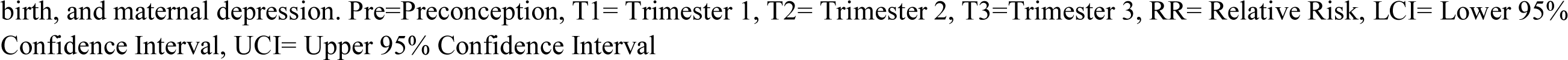
SMM risk across heat wave severity metrics (high-, moderate-, and low-intensity heat wave compared to no heat wave) for the warm season and summer season, South Carolina 1999-2017.

#### Air quality adjusted models

Poor air quality data were sparse in SC and resulted in the loss of 188,572 delivery records without a paired ozone measure) after merging in county-level air quality data in the larger cohort. Ozone adjusted full models revealed slightly attenuated adjusted RRs, but similar magnitudes of association between exposure to hot and cold extremes and SMM risks (Table S4).

#### Urban compared to rural SMM risks

Overall, the risk of a SMM event was higher in rural locations following cold exposure in T3 and for urban locations in T1 in response to a low vs high number of temperatures, but results were insignificant across most exposure windows and for hot ambient temperatures (Table S5). The risk of an SMM event in response to a low versus high number of very cold or cold temperatures during T3 was detected in both urban and rural settings, but higher SMM risks in response to cold extremes were observed among women in rural areas.

**Table 5.**
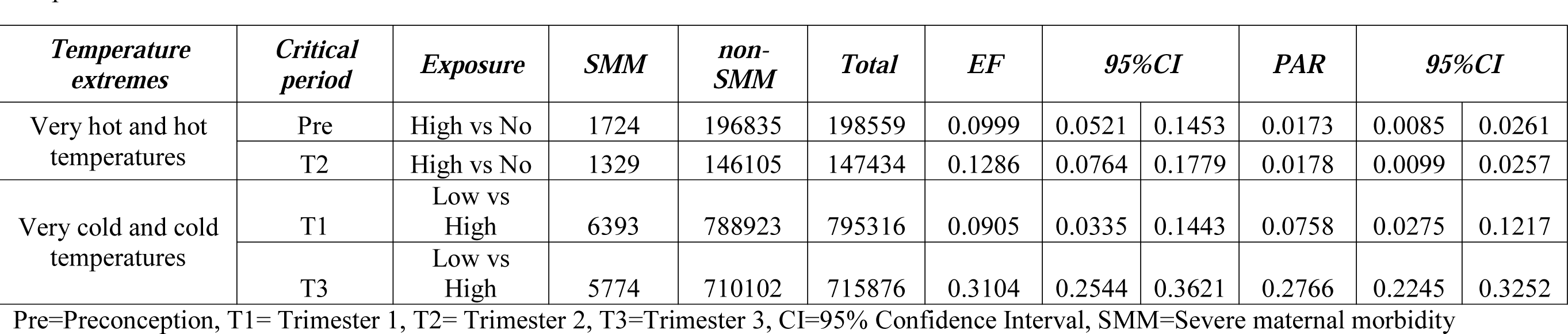
Excess fraction (EF), a proxy for attributable risk, and population attributable risk (PAR) of SMM due to exposure to hot and cold temperature extremes.

#### Advanced maternal age

In general, the risk of SMM following cold/very cold temperatures in T1 and T3 and very hot temperatures in the Pre period was higher for pregnant populations 35 years or younger (Table S5). However, we noted that women of advanced maternal age who were exposed to very hot or hot temperatures during T1 were 23% and 17% more likely to experience an SMM event, respectively (Table S6).

#### Preterm compared to Term SMM risks

Women with term SMM deliveries were more sensitive to cold exposure in T1 and T3 and women with preterm SMM deliveries were more susceptible to very hot temperatures in T2 (Table S6). For hot ambient temperature, results showed that women exposed to a high number of very hot days in T2 compared to no exposure were 20% more likely to experience a preterm SMM delivery.

#### Racialized and economic segregation

The risk of SMM following low compared to high exposure to cold temperatures in T3 was 1.45 times more likely for deliveries in low-income, black communities and 1.32 times more likely for deliveries in high-income, white communities (Table S8). A similar pattern was observed for very cold temperatures.

#### SMM risk attributable to temperature

Table 5 demonstrates the proportion of new SMM cases that are attributable to cold and hot temperature extremes (i.e., the excess fraction (EF)) and the proportion of new SMM cases in the total population of exposed and unexposed birthing people that are due to maternal exposure to thermal cold and hot extremes (i.e., population attributable risk). The EF for high vs no days of hot ambient temperature indicates that 9.9% and 12.9% of all SMM events in the high vs no exposure group are attributable to hot ambient temperature in the preconception and T2 periods, respectively. The PAR indicates that about 1.7% and 1.8% of all SMM events in the total population are attributable to heat exposure in the preconception and T2 periods, respectively.

For cold exposure, the EF revealed that 9.1% and 31.0% of all SMM events in the low compared to high number of cold days group are attributable to cold temperatures in T1 and T3, respectively. The PAR indicates that about 7.6% and 27.8% of all SMM events in the total population are attributable to cold exposure in the T1 and T3 periods, respectively.

## Discussion

The climate crisis poses important health threats for pregnant populations. The disproportionate exposure to climate-related changes in cold and hot temperature extremes may explain, in part, important racial differences in maternal health outcomes. At the time of publication, this study is the first to show an association between climate-sensitivities in extreme cold and hot ambient temperatures and increased maternal risk for ‘near miss’ events during critical windows of pregnancy. Results revealed an increase in SMM risk for pregnant individuals following unseasonably cold exposure during the first and third trimesters and exposure to hotter than average temperatures in the second trimester. We observed that maternal exposure to a low number of cold or very cold days, particularly in the third trimester, was associated with higher SMM risks. Conversely, sustained exposure to a high number of very hot days (i.e., consecutive number of 3 or more days), indicated by a heat wave, was associated with increased SMM risks in second trimester.

Findings showed important Black-white disparities in SMM risk following a high number of very cold days in the third trimester and heightened SMM risk for Black compared to white women following exposure to a small number of very hot days in the second trimester. The risk of SMM related to cold exposure in the third trimester was particularly high for younger pregnant populations (i.e., less than 35 years of age), those in rural areas, and for women residing in the most disadvantaged communities. Results identified important differences in cold temperature experience in T3 and SMM risk based on income inequality and structural racism.

Lastly, the risk of a term SMM delivery was higher in first trimester and third trimester following cold exposure and the risk of a preterm SMM delivery was higher in the second trimester following very hot temperatures. Findings suggest that maternal exposure to very hot or colder than average temperature extremes during critical periods of pregnancy may be a contributing environmental risk factor for SMM.

Cold ambient temperatures have been linked to poor birth outcomes in countries like Sweden (46), but have shown null results in US locations like California (47). One study demonstrated an increased risk for hypertensive disorders during pregnancy (i.e., eclampsia is one of the 21 disorders that comprise a SMM delivery) in the presence of exposure during the preconception period to very cold temperatures for pregnant women in China (22). A recent review examining seasonal variation in maternal disorders globally, like preeclampsia and eclampsia, highlighted a statistically significant relationship between maternal outcomes and lower ambient temperatures (48).

Cold temperatures have been associated with physiological reactivity among mammals influencing brown adipose tissue (49). In fact, one study showed that prenatal exposure to 18°C in mice initiated fetal fat brown fat storage; whereby, cold temperatures experienced during the third trimester may initiate cold-stress-related changes in the placenta (50). Cold stress in mice during the prenatal period has also been associated with negative impacts on a number of neuroimmune pathways, particularly as it pertains to the maternal hippocampus (51) and placental changes following late in pregnancy exposure (52). Vasoconstriction in the winter months has been identified as one possible explanation (53), while others have proposed environmental exposures encountered in the month of conception as important determinants for eclampsia onset (54). Some work has suggested that cold temperatures may increase exposure to passive second-hand smoking or infectious agents (55, 56) potentially increasing the risk of preeclampsia or hypertension in pregnant women (57). Animal models have laid the foundation for the need to prevent cold exposure and more targeted cold-reduction interventions later in pregnancy. However, the etiology linking colder temperatures during the preconception and prenatal periods is poorly understood and more research is needed to identify the multifactorial conditions leading up to a SMM delivery.

Our study found that Black compared to white birthing people were 63% and 70% more likely to experience a SMM delivery following exposure to a low and high number of very hot ambient temperature days in the second semester, respectively. We noted a similar SMM risk pattern association among Black compared to white pregnant populations in the first trimester. Our work parallels other studies highlighting Black compared to White mothers are more susceptible to high ambient temperatures, but prior studies have pointed to increased risk following heat exposure in the third trimester (2). Black women have higher rates of preterm hospitalization after hot temperature exposure in their first and third trimesters in Arizona, Washington, and New York (58) or higher preterm birth among Black birthing populations in the third trimester in Northern California (47). Black women may be more exposed to extreme heat due to their resident locations in areas with less tree canopy and limited access to mitigating measures (e.g., air conditioning) (59). But more research is needed to understand the individual-level and community-level factors that might be responsible for higher exposure to hotter ambient temperatures in or nearby Black maternal residences.

Early studies showed that prenatal exposure to hot ambient temperatures during the second and third trimesters was linked with a reduction in birth weight (60, 61). A recent review documented the maternal health effects of heat wave exposure and eclampsia, hypertensive disorders, uterine bleeding, and placental dysfunction (14). While epidemiologic evidence has shown that pregnant populations are at greater risk of heat stress (62), limited studies have explored the mechanisms underpinning pregnancy risk following heat exposure in the second trimester. One animal study showed that prolonged exposure to heat (35°C) during the second trimester was associated with the expression of heat shock response in placental tissue (63). High levels of heat shock proteins (HSP) --” a family of stress-induced proteins that are upregulated by a variety of stressors such as heat shock, steroids, infection, heavy metals” (64)-- are expressed in the placenta throughout pregnancy, but HSP 27 has a high pattern of expression in the second and third trimesters (65). Some studies have shown that high concentrations of HSP70 may be a marker for preterm delivery (66) and serum concentration of this protein declines throughout normal pregnancy development (67). Other studies have cited HSP70 as a potential biomarker for heat stress, in which patients experiencing heat-related illness expressed higher levels of this protein compared to a control group (68). Heat intolerance has been associated with cellular changes in gene expression, including increased production of HSP70 in humans (69, 70) and animal models (51, 71). While still limited, the current evidence base suggests that individuals with higher levels of HSP70 may be more susceptible to heat or respond maladaptively in the presence of excess heat exposure (68). Early evidence from animal models suggests that maternal exposure to ambient temperature extremes may perturb the gut microbiome during pregnancy with important implications for seasonal changes in thermoregulation (72).

### Clinical and research implications

A growing evidence-base is pointing towards the health harming effects of climate change, including exposure to extreme temperatures and weather events, for pregnant populations. Ambient temperature exposure is a modifiable risk factor for SMM. A priority area involves translating this research into changes in clinical practice and enhanced patient-centered care delivery, particularly in diverse care settings (e.g., low-income communities, minority communities, rural service areas). The integration of more education around avoiding extreme cold and hot ambient temperature, as well as other environmental risk factors like poor air quality, in preconception and prenatal care counseling is a necessary first step. Clinicians are trusted health messengers and can play a leading role in consulting with women on identifying and implementing preventive measures to reduce environmental hazards during these critical periods. In addition to raising awareness of environmental risk factors, clinicians can also advance national and state-level climate change policy (73).

Research on the influence of social-environmental drivers like residential segregation, poverty, and climate-related environmental exposures on maternal health disparities is lacking, and future climate change threatens recent maternal health progress (74). In turn, a renewed focus on the mechanistic link and pathways linking structural factors, like racism, with the climate crisis is needed to understand the full potential for increased maternal and child health disparities within a changing climate (75). For instance, racially segregated communities may be more vulnerable to climate stressors, like cold or hot ambient temperature extremes, due to structural factors like substandard housing that is too costly to heat or cool, low tree canopy, and a lack of political voice to motivate climate action and mitigation policy manifesting as geographically concentrated disadvantage (76). Research can be advanced through a multilevel framework to evaluate how area-level factors contribute to the etiologies of racial disparities in maternal and birth outcomes to identify key intervention opportunities at the clinical, community, and policy level.

Another critical research need is identifying a surveillance indicator to monitor and predict the maternal health impacts of climate change, identify vulnerable subgroups, and evaluate the efficacy of pregnancy interventions (77). National surveillance efforts are underway to monitor maternal health status, as well as to identity health system failures, preventable or modifiable risk factors, and intervention points to reduce the chronic health burden, medical care costs, lengthy hospital stays, and increased risks of maternal death. Our work highlights the potential use of SMM as a robust and credible health indicator of climate vulnerability that can be useful to health officials and policymakers in monitoring the effects of climate change on maternal health over time and evaluate progress toward adaptive strategies (78).

### Strengths and limitations

A key strength is that we included a comprehensive list of potential confounders, including maternal age, race/ethnicity, adequacy of prenatal care, obstetric conditions, and other neighborhood factors like urbanicity and racialized economic segregation. To further substantiate our results, we conducted multiple sensitivity analyses by including local air quality data, a known confounder, and the influence of low, moderate, or high intensity heat waves metric that accounts for local acclimatization. Selection bias was likely not an issue for our population-based approach of using hospital delivery discharges because the majority of births in SC occur in hospitals. Only a few studies have examined the effects of temperature exposure over vulnerable windows of pregnancy and even fewer have relied on a cohort design or have simultaneously examined both cold and heat-related temperatures (19, 20). An additional advantage of the retrospective cohort design involved the assessment of the temporal sequence of SMM risk and temperature exposure (79).

The following were limitations. First, we relied on an administrative dataset used for billing purposes. Due to coding errors, nondifferential misclassification may be present, underestimating the association between thermal temperature extremes and SMM. Temperature exposure assessment was based on in-situ station-based weather monitoring aggregated to maternal county residence and may have resulted in misclassification of temperature experience for individual women. Further, by relying on county-level temperature exposure estimates, estimates aggregated to this larger geographic unit may have reduced geographic variability and therefore lowered power to detect a true difference. Although hot and cold temperature exposure is typically ubiquitous for a given geographic area, emerging evidence from studies collecting individual experienced data suggest significant heterogeneity in temperature experience based on individual microclimates (80, 81). Lastly, other potential confounders, like ambient humidity exposure, BMI, or maternal behavioral activity patterns were not included due to a lack of data availability.

## Conclusions

To our knowledge, this is the first study to assess the relationship between prenatal exposure to temperature extremes and SMM risks in a Southern state using twenty years of data. Our findings suggest that maternal exposure to ambient temperature extremes is a modifiable risk factor for SMM. This study considered contextual social and environmental factors associated with increased SMM risks, such as residential segregation (a proxy for structural racism), residential poverty, and rural compared to urban differences. Surveillance of ‘near miss’events in the US is already underway and this research highlights the strong potential for the use of SMM as a valid indicator of the health impacts of climate change on at-risk birthing populations. More epidemiologic and policy focused research is needed to disentangle the complex relationship between structural factors and differential vulnerability to climate factors to identify preventive factors to minimize future climate-induced maternal health disparities.

## Data Availability Statement

Maternal health data used in this study are subject to a data use agreement and may not be shared.

## Data Availability

All data produced in the present work are contained in the manuscript

## Acknowledgements

We acknowledge Chris Finney and Sarah Crawford at the SC Revenue and Fiscal Affairs Office in the Health and Demographics Division for their technical expertise in providing data support.

## Author Contributions

JR designed and conceptualized the study; performed analysis; and lead the writing of the manuscript. SS contributed to data extraction/manipulation and the writing of the paper. MS contributed to the methods development, analysis, and writing of the manuscript.

## Ethical Approval

Approval to use these data and conduct this secondary analysis was approved by the North Carolina State University Institutional Review Board (IRB# 24297).

## Acknowledgements

This work was supported in part by the National Oceanic and Atmospheric Administration (NOAA) through the Cooperative Institute for Satellite Earth System Studies under Cooperative Agreement NA19NES4320002 and the NOAA Regional Integrated Sciences and Assessments (RISA) program NA21OAR4310312.

