## Supplemental Materials for "Contribution of prenatal exposure to ambient temperature extremes and severe maternal morbidity: A retrospective Southern birth cohort"

**Supplemental Tables**

Table S1. Description of maternal health outcomes used in the analysis.

| *Maternal outcome* | *ICD-9 code(s)* |
| --- | --- |
| Severe maternal morbidity (SMM) | 1. Acute myocardial infarction DX 410.xx; 2. Aneurysm DX 441.xx; 3. Acute renal failure DX 584.5, 584.6, 584.7, 584.8, 584.9, 669.3x; 4. Adult respiratory distress syndrome DX 518.5x, 518.81 518.82 518.84, 799.1; 5. Amniotic fluid embolism DX 673.1x; 6. Cardiac arrest/ventricular fibrillation DX 427.41, 427.42, 427.5; 7. Conversion of cardiac rhythm PR 99.6x; 8. Disseminated intravascular coagulation DX 286.6, 286.9, 666.3x; 9. Eclampsia DX 642.6x; 10. Heart failure/arrest during surgery or procedure DX 997.1; 11. Puerperal cerebrovascular disorders DX 430.xx, 431.xx, 432.xx, 433.xx, 434.xx, 436xx, 437.xx, 671.5x, 674.0x, 997.02; 12. Pulmonary edema / Acute heart failure DX 518.4, 428.1, 428.0, 428.21, 428.23, 428.31, 428.33, 428.41, 428.43; 13. Severe anesthesia complications DX 668.0x, 668.1x, 668.2x; 14. Sepsis DX 038.xx, 995.91, 995.92, 670.2x; 15. Shock DX 669.1x, 785.5x, 995.0, 995.4, 998.0x; 16. Sickle cell disease with crisis DX 282.42, 282.62, 282.64, 282.6; 17. Air and thrombotic embolism DX 415.1x, 673.0x, 673.2x 673.3x, 673.8x; 18. Blood products transfusion PR 99.0x; 19. Hysterectomy PR 68.3x-68.9x; 20. Temporary tracheostomy PR 31.1; 21.Ventilation PR 93.90, 96.01, 96.02, 96.03, 96.05 |

ICD-9 = International Classification of Disease, Ninth Revision; DX = Diagnosis code; PR = Procedure code

Table S2. Distribution of average temperature across critical periods.

|  |  | Mean±SD | 50th | 75th | 90th | 95th | 99th | IQR | Range |
| --- | --- | --- | --- | --- | --- | --- | --- | --- | --- |
| Pre | Very cold | 4.7 (7.2) | 0 | 7 | 16 | 21 | 29 | 7 | (0, 41) |
|  | Cold | 4.6 (6.2) | 0 | 8 | 14 | 17 | 23 | 8 | (0, 35) |
|  | Very hot | 4.5 (8.4) | 0 | 5 | 17 | 24 | 36 | 5 | (0, 55) |
|  | Hot | 4.2 (6.0) | 0 | 8 | 14 | 17 | 22 | 8 | (0, 32) |
| T1 | Very cold | 5.3 (7.7) | 0 | 9 | 18 | 22 | 30 | 9 | (0, 41) |
|  | Cold | 5.2 (6.6) | 0 | 10 | 15 | 19 | 24 | 10 | (0, 36) |
|  | Very hot | 4.8 (8.7) | 0 | 6 | 18 | 25 | 37 | 6 | (0, 55) |
|  | Hot | 4.4 (6.3) | 0 | 8 | 15 | 17 | 23 | 8 | (0, 33) |
| T2 | Very cold | 4.6 (7.2) | 0 | 7 | 16 | 20 | 29 | 7 | (0, 41) |
|  | Cold | 4.6 (6.2) | 0 | 8 | 14 | 17 | 23 | 8 | (0, 35) |
|  | Very hot | 4.4 (8.3) | 0 | 5 | 17 | 24 | 37 | 5 | (0, 55) |
|  | Hot | 4.2 (6.0) | 0 | 8 | 14 | 17 | 22 | 8 | (0, 32) |
| T3 | Very cold | 4.1 (6.8) | 0 | 6 | 15 | 19 | 29 | 6 | (0, 41) |
|  | Cold | 4.0 (5.8) | 0 | 7 | 13 | 16 | 21 | 7 | (0, 36) |
|  | Very hot | 4.2 (8.0) | 0 | 5 | 16 | 23 | 35 | 5 | (0. 55) |
|  | Hot | 3.9 (5.7) | 0 | 7 | 13 | 16 | 22 | 7 | (0, 33) |

Note: Pre=Preconception, T1= Trimester 1, T2= Trimester 2, T3=Trimester 3, SD= Standard deviation, IQR=Interquartile range

Table S3. Very hot and hot temperature exposure for T2 and T3 restricted to the summer season.

|  |  |  | Adjusted models | | |
| --- | --- | --- | --- | --- | --- |
| T2 | **Very hot (95th percentile)** | | **RR** | **LCI** | **UCI** |
|  | low vs high exposure | | 0.76 | 0.44 | 1.32 |
|  | low vs no exposure | | 1.55 | 1.24 | 1.93 |
|  | high vs no exposure | | 2.03 | 1.21 | 3.40 |
|  | **Hot (90th percentile)** | | **RR** | **LCI** | **UCI** |
|  | low vs high exposure | | 0.66 | 0.38 | 1.15 |
|  | low vs no exposure | | 1.41 | 1.17 | 1.69 |
|  | high vs no exposure | | 2.12 | 1.25 | 3.62 |
| T3 | **Very hot (95th percentile)** | | **RR** | **LCI** | **UCI** |
|  | low vs high exposure | | 1.14 | 0.98 | 1.34 |
|  | low vs no exposure | | 0.96 | 0.84 | 1.10 |
|  | high vs no exposure | | 0.84 | 0.70 | 1.00 |
|  | **Hot (90th percentile)** | | **RR** | **LCI** | **UCI** |
|  | low vs high exposure | | 1.01 | 0.88 | 1.17 |
|  | low vs no exposure | | 0.95 | 0.80 | 1.12 |
|  | high vs no exposure | | 0.93 | 0.77 | 1.13 |

Note: All estimates from GEE models adjusted for maternal age, race/ethnicity, insurance payor, preexisting diabetes, chronic hypertension, obesity, chronic renal disease, abnormal presentation, prior cesarean, parity, placental abnormality, adequacy of prenatal care, smoking, year of birth, and maternal depression. T2= Trimester 2, T3=Trimester 3, RR= Relative Risk, LCI= Lower 95%

Confidence Interval, UCI= Upper 95% Confidence Interval

Table S4. Full temperature models adjusted^a^ for ozone.

| T1 | **Cold (10th)** | **RR** | **LCI** | **UCI** |
| --- | --- | --- | --- | --- |
|  | low vs high exposure | 0.90 | 0.83 | 0.98 |
|  | low vs no exposure | 0.96 | 0.90 | 1.01 |
|  | high vs no exposure | 1.06 | 0.97 | 1.15 |
|  | **Very cold (5th)** | **RR** | **LCI** | **UCI** |
|  | low vs high exposure | 1.08 | 0.94 | 1.24 |
|  | low vs no exposure | 0.97 | 0.92 | 1.04 |
|  | high vs no exposure | 0.90 | 0.78 | 1.04 |
| T2 | **Hot (90th)** | **RR** | **LCI** | **UCI** |
|  | low vs high exposure | 1.11 | 1.02 | 1.20 |
|  | low vs no exposure | 1.02 | 0.96 | 1.08 |
|  | high vs no exposure | 0.92 | 0.84 | 1.00 |
|  | **Very hot (95th)** | **RR** | **LCI** | **UCI** |
|  | low vs high exposure | 0.93 | 0.83 | 1.05 |
|  | low vs no exposure | 1.00 | 0.94 | 1.06 |
|  | high vs no exposure | 1.07 | 0.96 | 1.20 |
| T3 | **Cold (10th)** | **RR** | **LCI** | **UCI** |
|  | low vs high exposure | **1.33** | **1.19** | **1.49** |
|  | low vs no exposure | 0.99 | 0.93 | 1.04 |
|  | high vs no exposure | 0.74 | 0.66 | 0.83 |
|  | **Very cold (5th)** | **RR** | **LCI** | **UCI** |
|  | low vs high exposure | **1.37** | **1.13** | **1.66** |
|  | low vs no exposure | 1.01 | 0.94 | 1.09 |
|  | high vs no exposure | 0.74 | 0.61 | 0.89 |

Note: All estimates from GEE models adjusted for maternal age, race/ethnicity, insurance payor, preexisting diabetes, chronic hypertension, obesity, chronic renal disease, abnormal presentation, prior cesarean, parity, placental abnormality, adequacy of prenatal care, smoking, year of birth, and maternal depression. ^a^Only significant adjusted models were included in the results. Pre=Preconception, T1= Trimester 1, T2= Trimester 2, T3=Trimester 3,RR= Relative Risk, LCI= Lower 95% Confidence Interval, UCI= Upper 95% Confidence Interval.

Table S5. Adjusted models of SMM risk for cold and hot temperature extremes across pregnancy periods for urban and rural maternal residence and maternal age.

|  |  |  | **Rural vs Urban** | | | | | | | | | | | | **Maternal Age** | | | | | | | | | | | |
| --- | --- | --- | --- | --- | --- | --- | --- | --- | --- | --- | --- | --- | --- | --- | --- | --- | --- | --- | --- | --- | --- | --- | --- | --- | --- | --- |
|  |  |  | **Urban** | | | **Rural** | | | **Urban** | | | **Rural** | | | **Age < 35 yrs** | | | **Age ≥35 yrs** | | | **Age < 35 yrs** | | | **Age ≥35 yrs** | | |
|  | **Exposure level** | | **RR** | **95% CI** | | **RR** | **95% CI** | | **RR** | **95% CI** | | **RR** | **95% CI** | | **RR** | **95% CI** | | **RR** | **95% CI** | | **RR** | **95% CI** | | **RR** | **95% CI** | |
| **Pre** |  |  | **Very Cold (< 5th)** | | | | | | **Very Hot (< 95th)** | | | | | | **Very Cold (< 5th)** | | | | | | **Very Hot (< 95th)** | | | | | |
|  | low vs high exposure | | 1.08 | 0.96 | 1.21 | 0.97 | 0.78 | 1.19 | 0.92 | 0.82 | 1.03 | 0.96 | 0.78 | 1.18 | 1.06 | 0.95 | 1.18 | 1.02 | 0.80 | 1.31 | 0.91 | 0.82 | 1.01 | 1.02 | 0.80 | 1.30 |
|  | low vs no exposure | | 0.98 | 0.92 | 1.05 | 0.96 | 0.84 | 1.10 | 1.00 | 0.93 | 1.08 | 1.04 | 0.91 | 1.20 | 0.97 | 0.91 | 1.04 | 1.00 | 0.87 | 1.17 | 1.01 | 0.94 | 1.09 | 1.01 | 0.86 | 1.18 |
|  | high vs no exposure | | 0.91 | 0.81 | 1.02 | 1.00 | 0.81 | 1.22 | 1.09 | 0.99 | 1.21 | 1.09 | 0.90 | 1.32 | 0.92 | 0.83 | 1.02 | 0.98 | 0.78 | 1.24 | 1.11 | 1.01 | 1.23 | 0.99 | 0.79 | 1.24 |
|  |  |  | **Cold (<10th)** | | | | | | **Hot (<90th)** | | | | | | **Cold (<10th)** | | | | | | **Hot (<90th)** | | | | | |
|  | low vs high exposure | | 1.06 | 0.95 | 1.18 | 1.03 | 0.84 | 1.26 | 0.95 | 0.85 | 1.06 | 0.87 | 0.71 | 1.07 | 1.07 | 0.97 | 1.20 | 0.94 | 0.75 | 1.17 | 0.94 | 0.84 | 1.04 | 0.90 | 0.71 | 1.13 |
|  | low vs no exposure | | 0.98 | 0.92 | 1.05 | 0.94 | 0.82 | 1.07 | 1.01 | 0.94 | 1.09 | 1.01 | 0.89 | 1.15 | 0.97 | 0.90 | 1.03 | 0.99 | 0.85 | 1.15 | 1.02 | 0.95 | 1.09 | 0.98 | 0.84 | 1.13 |
|  | high vs no exposure | | 0.93 | 0.84 | 1.03 | 0.91 | 0.75 | 1.11 | 1.07 | 0.96 | 1.19 | 1.16 | 0.96 | 1.41 | 0.90 | 0.81 | 1.00 | 1.06 | 0.85 | 1.32 | 1.09 | 0.98 | 1.21 | 1.09 | 0.88 | 1.36 |
| **T1** |  |  | **Very Cold (< 5th)** | | | | | | **Very Hot (< 95th)** | | | | | | **Very Cold (< 5th)** | | | | | | **Very Hot (< 95th)** | | | | | |
|  | low vs high exposure | | 1.18 | 1.04 | 1.33 | 0.99 | 0.80 | 1.23 | 0.96 | 0.86 | 1.07 | 1.13 | 0.91 | 1.41 | 1.15 | 1.03 | 1.29 | 1.04 | 0.80 | 1.35 | 0.99 | 0.88 | 1.10 | 1.03 | 0.82 | 1.30 |
|  | low vs no exposure | | 1.03 | 0.96 | 1.10 | 0.98 | 0.87 | 1.12 | 1.01 | 0.94 | 1.09 | 1.05 | 0.92 | 1.21 | 1.05 | 0.99 | 1.13 | 0.86 | 0.74 | 1.00 | 0.98 | 0.91 | 1.05 | 1.23 | 1.05 | 1.43 |
|  | high vs no exposure | | 0.87 | 0.78 | 0.98 | 0.99 | 0.80 | 1.22 | 1.05 | 0.95 | 1.17 | 0.93 | 0.76 | 1.14 | 0.91 | 0.82 | 1.02 | 0.83 | 0.65 | 1.06 | 0.99 | 0.90 | 1.10 | 1.19 | 0.96 | 1.47 |
|  |  |  | **Cold (<10th)** | | | | | | **Hot (<90th)** | | | | | | **Cold (<10th)** | | | | | | **Hot (<90th)** | | | | | |
|  | low vs high exposure | | 1.11 | 1.00 | 1.24 | 1.12 | 0.91 | 1.37 | 1.03 | 0.92 | 1.15 | 1.11 | 0.89 | 1.38 | 1.13 | 1.02 | 1.26 | 1.04 | 0.82 | 1.32 | 1.05 | 0.94 | 1.18 | 1.01 | 0.80 | 1.27 |
|  | low vs no exposure | | 1.02 | 0.95 | 1.09 | 1.03 | 0.90 | 1.17 | 1.02 | 0.96 | 1.10 | 1.03 | 0.90 | 1.17 | 1.05 | 0.99 | 1.13 | 0.88 | 0.76 | 1.02 | 1.00 | 0.93 | 1.07 | 1.17 | 1.01 | 1.35 |
|  | high vs no exposure | | 0.92 | 0.82 | 1.02 | 0.92 | 0.75 | 1.12 | 1.00 | 0.89 | 1.12 | 0.93 | 0.75 | 1.15 | 0.93 | 0.84 | 1.04 | 0.85 | 0.67 | 1.07 | 0.95 | 0.85 | 1.06 | 1.16 | 0.92 | 1.46 |
| **T2** |  |  | **Very Cold (< 5th)** | | | | | | **Very Hot (< 95th)** | | | | | | **Very Cold (< 5th)** | | | | | | **Very Hot (< 95th)** | | | | | |
|  | low vs high exposure | | 1.02 | 0.91 | 1.14 | 1.00 | 0.81 | 1.23 | 0.91 | 0.81 | 1.02 | 1.00 | 0.81 | 1.24 | 0.99 | 0.88 | 1.10 | 1.19 | 0.92 | 1.54 | 0.93 | 0.83 | 1.04 | 0.91 | 0.72 | 1.15 |
|  | low vs no exposure | | 1.01 | 0.94 | 1.09 | 1.07 | 0.94 | 1.22 | 0.97 | 0.90 | 1.05 | 0.98 | 0.86 | 1.13 | 1.02 | 0.95 | 1.09 | 1.04 | 0.90 | 1.21 | 0.97 | 0.90 | 1.04 | 1.01 | 0.87 | 1.19 |
|  | high vs no exposure | | 0.99 | 0.89 | 1.11 | 1.07 | 0.87 | 1.31 | 1.07 | 0.97 | 1.19 | 0.99 | 0.81 | 1.20 | 1.04 | 0.93 | 1.15 | 0.87 | 0.68 | 1.12 | 1.04 | 0.94 | 1.15 | 1.12 | 0.90 | 1.39 |
|  |  |  | **Cold (<10th)** | | | | | | **Hot (<90th)** | | | | | | **Cold (<10th)** | | | | | | **Hot (<90th)** | | | | | |
|  | low vs high exposure | | 1.01 | 0.91 | 1.13 | 0.93 | 0.77 | 1.14 | 0.90 | 0.81 | 1.00 | 0.93 | 0.76 | 1.15 | 0.99 | 0.89 | 1.09 | 1.02 | 0.81 | 1.30 | 0.90 | 0.81 | 1.00 | 0.97 | 0.77 | 1.22 |
|  | low vs no exposure | | 1.01 | 0.95 | 1.09 | 0.98 | 0.86 | 1.12 | 0.95 | 0.89 | 1.02 | 0.96 | 0.84 | 1.09 | 1.01 | 0.94 | 1.08 | 0.99 | 0.86 | 1.15 | 0.96 | 0.89 | 1.02 | 0.94 | 0.81 | 1.09 |
|  | high vs no exposure | | 1.00 | 0.90 | 1.11 | 1.05 | 0.86 | 1.27 | 1.06 | 0.95 | 1.17 | 1.03 | 0.84 | 1.25 | 1.02 | 0.92 | 1.13 | 0.97 | 0.77 | 1.22 | 1.06 | 0.96 | 1.18 | 0.97 | 0.78 | 1.21 |
| **T3** |  |  | **Very Cold (< 5th)** | | | | | | **Very Hot (< 95th)** | | | | | | **Very Cold (< 5th)** | | | | | | **Very Hot (< 95th)** | | | | | |
|  | low vs high exposure | | 1.21 | 1.07 | 1.37 | 1.13 | 0.90 | 1.43 | 1.06 | 0.94 | 1.20 | 0.95 | 0.76 | 1.18 | 1.25 | 1.11 | 1.42 | 0.95 | 0.74 | 1.21 | 1.03 | 0.92 | 1.16 | 1.04 | 0.80 | 1.36 |
|  | low vs no exposure | | 1.02 | 0.95 | 1.09 | 0.96 | 0.84 | 1.09 | 0.94 | 0.87 | 1.01 | 0.93 | 0.81 | 1.07 | 0.99 | 0.92 | 1.06 | 1.10 | 0.95 | 1.28 | 0.95 | 0.88 | 1.02 | 0.88 | 0.75 | 1.03 |
|  | high vs no exposure | | 0.84 | 0.75 | 0.95 | 0.84 | 0.68 | 1.06 | 0.88 | 0.79 | 0.99 | 0.98 | 0.80 | 1.21 | 0.79 | 0.70 | 0.88 | 1.16 | 0.92 | 1.47 | 0.91 | 0.82 | 1.02 | 0.85 | 0.66 | 1.08 |
|  |  |  | **Cold (<10th)** | | | | | | **Hot (<90th)** | | | | | | **Cold (<10th)** | | | | | | **Hot (<90th)** | | | | | |
|  | low vs high exposure | | 1.29 | 1.14 | 1.45 | 1.36 | 1.08 | 1.71 | 1.04 | 0.92 | 1.16 | 1.08 | 0.86 | 1.34 | 1.35 | 1.20 | 1.52 | 1.10 | 0.87 | 1.41 | 1.04 | 0.93 | 1.16 | 1.06 | 0.82 | 1.37 |
|  | low vs no exposure | | 1.03 | 0.96 | 1.10 | 1.01 | 0.89 | 1.15 | 0.94 | 0.88 | 1.01 | 0.97 | 0.85 | 1.10 | 1.01 | 0.94 | 1.08 | 1.12 | 0.97 | 1.29 | 0.97 | 0.91 | 1.04 | 0.84 | 0.73 | 0.98 |
|  | high vs no exposure | | 0.80 | 0.71 | 0.90 | 0.74 | 0.59 | 0.93 | 0.91 | 0.82 | 1.02 | 0.90 | 0.73 | 1.11 | 0.75 | 0.67 | 0.84 | 1.01 | 0.80 | 1.28 | 0.93 | 0.84 | 1.04 | 0.80 | 0.62 | 1.01 |

Note: All estimates from GEE models adjusted for maternal age, race/ethnicity, insurance payor, preexisting diabetes, chronic hypertension, obesity, chronic renal disease, abnormal presentation, prior cesarean, parity, placental abnormality, adequacy of prenatal care, smoking, year of birth, and maternal depression. Pre=Preconception, T1= Trimester 1, T2= Trimester 2, T3=Trimester 3,RR= Relative Risk, LCI= Lower 95% Confidence Interval, UCI= Upper 95% Confidence Interval.

Table S6. Adjusted models of SMM risk for cold and hot temperature extremes across pregnancy periods for preterm and term SMM.

|  | COLD EXPOSURE | *Preterm SMM* | | | *Term SMM* | | | HEAT EXPOSURE | *Preterm SMM* | | | *Term SMM* | | |
| --- | --- | --- | --- | --- | --- | --- | --- | --- | --- | --- | --- | --- | --- | --- |
| Pre | **Very cold (5th )** | **RR** | **LCI** | **UCI** | **RR** | **LCI** | **UCI** | **Very hot (95th )** | **RR** | **LCI** | **UCI** | **RR** | **LCI** | **UCI** |
|  | low vs high exposure | 0.95 | 0.78 | 1.15 | 1.10 | 0.98 | 1.24 | low vs high exposure | 0.88 | 0.72 | 1.07 | 0.94 | 0.84 | 1.06 |
|  | low vs no exposure | 1.01 | 0.89 | 1.14 | 0.97 | 0.90 | 1.04 | low vs no exposure | 0.97 | 0.85 | 1.10 | 1.03 | 0.95 | 1.11 |
|  | high vs no exposure | 1.06 | 0.89 | 1.28 | 0.88 | 0.78 | 0.99 | high vs no exposure | 1.10 | 0.92 | 1.31 | 1.09 | 0.98 | 1.21 |
|  | **Cold (10th )** | **RR** | **LCI** | **UCI** | **RR** | **LCI** | **UCI** | **Hot (90th )** | **RR** | **LCI** | **UCI** | **RR** | **LCI** | **UCI** |
|  | low vs high exposure | 0.92 | 0.77 | 1.10 | 1.10 | 0.98 | 1.23 | low vs high exposure | 0.91 | 0.75 | 1.10 | 0.94 | 0.84 | 1.05 |
|  | low vs no exposure | 0.98 | 0.87 | 1.11 | 0.97 | 0.90 | 1.04 | low vs no exposure | 0.95 | 0.84 | 1.07 | 1.04 | 0.96 | 1.11 |
|  | high vs no exposure | 1.07 | 0.89 | 1.27 | 0.88 | 0.79 | 0.98 | high vs no exposure | 1.05 | 0.87 | 1.26 | 1.10 | 0.99 | 1.23 |
| T1 | **Very cold (5th )** | **RR** | **LCI** | **UCI** | **RR** | **LCI** | **UCI** | **Very hot (95th )** | **RR** | **LCI** | **UCI** | **RR** | **LCI** | **UCI** |
|  | low vs high exposure | 1.09 | 0.88 | 1.34 | 1.15 | 1.02 | 1.30 | low vs high exposure | 1.07 | 0.87 | 1.31 | 0.97 | 0.86 | 1.09 |
|  | low vs no exposure | 0.94 | 0.83 | 1.06 | 1.05 | 0.98 | 1.12 | low vs no exposure | 1.08 | 0.95 | 1.23 | 1.00 | 0.93 | 1.08 |
|  | high vs no exposure | 0.86 | 0.71 | 1.06 | 0.91 | 0.81 | 1.02 | high vs no exposure | 1.01 | 0.84 | 1.22 | 1.03 | 0.93 | 1.15 |
|  | **Cold (10th )** | **RR** | **LCI** | **UCI** | **RR** | **LCI** | **UCI** | **Hot (90th )** | **RR** | **LCI** | **UCI** | **RR** | **LCI** | **UCI** |
|  | low vs high exposure | 1.19 | 0.98 | 1.44 | 1.09 | 0.98 | 1.22 | low vs high exposure | 1.10 | 0.90 | 1.34 | 1.02 | 0.91 | 1.15 |
|  | low vs no exposure | 1.00 | 0.89 | 1.12 | 1.03 | 0.96 | 1.10 | low vs no exposure | 1.12 | 0.99 | 1.26 | 0.99 | 0.93 | 1.07 |
|  | high vs no exposure | 0.84 | 0.69 | 1.02 | 0.94 | 0.84 | 1.05 | high vs no exposure | 1.02 | 0.84 | 1.25 | 0.97 | 0.87 | 1.09 |
| T2 | **Very cold (5th )** | **RR** | **LCI** | **UCI** | **RR** | **LCI** | **UCI** | **Very hot (95th )** | **RR** | **LCI** | **UCI** | **RR** | **LCI** | **UCI** |
|  | low vs high exposure | 1.10 | 0.90 | 1.35 | 0.99 | 0.89 | 1.11 | low vs high exposure | 0.81 | 0.67 | 0.98 | 0.98 | 0.87 | 1.10 |
|  | low vs no exposure | 0.99 | 0.88 | 1.12 | 1.04 | 0.97 | 1.11 | low vs no exposure | 0.98 | 0.86 | 1.11 | 0.98 | 0.90 | 1.05 |
|  | high vs no exposure | 0.90 | 0.74 | 1.10 | 1.05 | 0.94 | 1.17 | high vs no exposure | 1.20 | 1.02 | 1.43 | 1.00 | 0.89 | 1.11 |
|  | **Cold (10th )** | **RR** | **LCI** | **UCI** | **RR** | **LCI** | **UCI** | **Hot (90th )** | **RR** | **LCI** | **UCI** | **RR** | **LCI** | **UCI** |
|  | low vs high exposure | 1.03 | 0.85 | 1.25 | 0.98 | 0.88 | 1.09 | low vs high exposure | 0.89 | 0.74 | 1.07 | 0.92 | 0.82 | 1.02 |
|  | low vs no exposure | 0.99 | 0.88 | 1.12 | 1.01 | 0.95 | 1.09 | low vs no exposure | 0.99 | 0.88 | 1.12 | 0.94 | 0.87 | 1.01 |
|  | high vs no exposure | 0.96 | 0.80 | 1.16 | 1.03 | 0.93 | 1.15 | high vs no exposure | 1.11 | 0.93 | 1.33 | 1.03 | 0.92 | 1.14 |
| T3 | **Very cold (5th )** | **RR** | **LCI** | **UCI** | **RR** | **LCI** | **UCI** | **Very hot (95th )** | **RR** | **LCI** | **UCI** | **RR** | **LCI** | **UCI** |
|  | low vs high exposure | 1.10 | 0.81 | 1.47 | 1.13 | 1.00 | 1.27 | low vs high exposure | 1.09 | 0.82 | 1.45 | 0.98 | 0.87 | 1.10 |
|  | low vs no exposure | 0.99 | 0.87 | 1.12 | 1.02 | 0.95 | 1.10 | low vs no exposure | 0.84 | 0.74 | 0.96 | 0.98 | 0.91 | 1.05 |
|  | high vs no exposure | 0.90 | 0.68 | 1.20 | 0.91 | 0.81 | 1.01 | high vs no exposure | 0.77 | 0.59 | 1.01 | 1.00 | 0.90 | 1.11 |
|  | **Cold (10th )** | **RR** | **LCI** | **UCI** | **RR** | **LCI** | **UCI** | **Hot (90th )** | **RR** | **LCI** | **UCI** | **RR** | **LCI** | **UCI** |
|  | low vs high exposure | 1.20 | 0.87 | 1.64 | 1.21 | 1.08 | 1.36 | low vs high exposure | 1.20 | 0.88 | 1.63 | 0.98 | 0.87 | 1.10 |
|  | low vs no exposure | 1.00 | 0.89 | 1.13 | 1.04 | 0.97 | 1.11 | low vs no exposure | 0.89 | 0.79 | 1.00 | 0.98 | 0.91 | 1.05 |
|  | high vs no exposure | 0.84 | 0.61 | 1.15 | 0.86 | 0.77 | 0.96 | high vs no exposure | 0.74 | 0.55 | 1.01 | 1.00 | 0.90 | 1.11 |

Note: All estimates from GEE models adjusted for maternal age, race/ethnicity, insurance payor, preexisting diabetes, chronic hypertension, obesity, chronic renal disease, abnormal presentation, prior cesarean, parity, placental abnormality, adequacy of prenatal care, smoking, year of birth, and maternal depression. Pre=Preconception, T1= Trimester 1, T2= Trimester 2, T3=Trimester 3,RR= Relative Risk, LCI= Lower 95% Confidence Interval, UCI= Upper 95% Confidence Interval.

Table S7. Adjusted models of SMM risk for cold and hot temperature extremes across pregnancy periods for racially and economically segregated communities.

|  |  |  |  | **Very cold (<5th)** | | | **Cold (<10th)** | | | **Very hot (>95th)** | | | **Hot (<90th)** | | |
| --- | --- | --- | --- | --- | --- | --- | --- | --- | --- | --- | --- | --- | --- | --- | --- |
|  | **ICE** | **Exposure** | | **RR** | **95%CI** | | **RR** | **95%CI** | | **RR** | **95%CI** | | **RR** | **95%CI** | |
| Pre | Q1: Most deprived | Low vs high | | 0.93 | 0.78 | 1.12 | 1.00 | 0.84 | 1.19 | 0.93 | 0.77 | 1.12 | 1.03 | 0.86 | 1.25 |
|  |  | Low vs no exposure | | 0.95 | 0.84 | 1.06 | 0.99 | 0.89 | 1.12 | 0.95 | 0.84 | 1.07 | 1.01 | 0.90 | 1.14 |
|  |  | High vs no exposure | | 1.01 | 0.85 | 1.21 | 0.99 | 0.84 | 1.18 | 1.02 | 0.86 | 1.22 | 0.98 | 0.82 | 1.17 |
|  | Q2 | Low vs high | | 1.02 | 0.85 | 1.22 | 0.97 | 0.82 | 1.15 | 0.94 | 0.79 | 1.12 | 0.93 | 0.78 | 1.10 |
|  |  | Low vs no exposure | | 0.96 | 0.86 | 1.07 | 0.93 | 0.83 | 1.04 | 1.07 | 0.95 | 1.21 | 1.04 | 0.93 | 1.16 |
|  |  | High vs no exposure | | 0.94 | 0.79 | 1.12 | 0.96 | 0.81 | 1.13 | 1.14 | 0.97 | 1.34 | 1.12 | 0.95 | 1.33 |
|  | Q3: Least deprived | Low vs high | | 1.18 | 1.00 | 1.40 | 1.16 | 0.99 | 1.36 | 0.92 | 0.79 | 1.08 | 0.86 | 0.74 | 1.00 |
|  |  | Low vs no exposure | | 1.02 | 0.93 | 1.13 | 0.99 | 0.90 | 1.09 | 1.02 | 0.91 | 1.13 | 0.98 | 0.89 | 1.08 |
|  |  | High vs no exposure | | 0.87 | 0.74 | 1.02 | 0.85 | 0.73 | 1.00 | 1.10 | 0.96 | 1.27 | 1.13 | 0.98 | 1.31 |
| T1 | Q1: Most deprived | Low vs high | | 1.08 | 0.86 | 1.36 | 1.05 | 0.88 | 1.25 | 1.14 | 0.87 | 1.49 | 1.08 | 0.89 | 1.31 |
|  |  | Low vs no exposure | | 1.02 | 0.91 | 1.14 | 1.02 | 0.91 | 1.14 | 0.99 | 0.88 | 1.11 | 0.99 | 0.89 | 1.12 |
|  | Q2 | Low vs high | | 1.00 | 0.81 | 1.24 | 1.04 | 0.88 | 1.24 | 1.18 | 0.92 | 1.52 | 1.10 | 0.92 | 1.32 |
|  |  | Low vs no exposure | | 0.92 | 0.83 | 1.02 | 0.94 | 0.84 | 1.05 | 1.11 | 1.00 | 1.24 | 1.13 | 1.02 | 1.26 |
|  |  | High vs no exposure | | 0.92 | 0.74 | 1.14 | 0.90 | 0.76 | 1.07 | 0.94 | 0.73 | 1.20 | 1.03 | 0.86 | 1.23 |
|  | Q3: Least deprived | Low vs high | | 1.24 | 1.01 | 1.53 | 1.22 | 1.04 | 1.42 | 0.90 | 0.73 | 1.12 | 0.97 | 0.83 | 1.14 |
|  |  | Low vs no exposure | | 1.07 | 0.98 | 1.18 | 1.08 | 0.98 | 1.19 | 0.99 | 0.90 | 1.09 | 0.97 | 0.88 | 1.07 |
|  |  | High vs no exposure | | 0.86 | 0.70 | 1.06 | 0.89 | 0.76 | 1.04 | 1.10 | 0.89 | 1.35 | 1.00 | 0.85 | 1.16 |
| T2 | Q1: Most deprived | Low vs high | | 1.12 | 0.86 | 1.47 | 1.03 | 0.85 | 1.24 | 0.99 | 0.78 | 1.26 | 0.89 | 0.75 | 1.07 |
|  |  | Low vs no exposure | | 1.01 | 0.90 | 1.12 | 0.99 | 0.88 | 1.11 | 1.00 | 0.89 | 1.12 | 0.97 | 0.86 | 1.09 |
|  |  | High vs no exposure | | 0.89 | 0.69 | 1.16 | 0.97 | 0.80 | 1.17 | 1.01 | 0.80 | 1.27 | 1.08 | 0.91 | 1.28 |
|  | Q2 | Low vs high | | 0.98 | 0.77 | 1.25 | 1.01 | 0.84 | 1.21 | 0.78 | 0.63 | 0.96 | 0.86 | 0.72 | 1.02 |
|  |  | Low vs no exposure | | 1.04 | 0.93 | 1.15 | 1.02 | 0.92 | 1.14 | 0.95 | 0.85 | 1.06 | 0.90 | 0.81 | 1.01 |
|  |  | High vs no exposure | | 1.06 | 0.83 | 1.34 | 1.01 | 0.85 | 1.22 | 1.22 | 1.00 | 1.49 | 1.05 | 0.90 | 1.24 |
|  | Q3: Least deprived | Low vs high | | 1.23 | 0.97 | 1.56 | 0.98 | 0.83 | 1.15 | 0.89 | 0.73 | 1.09 | 0.95 | 0.81 | 1.11 |
|  |  | Low vs no exposure | | 1.04 | 0.94 | 1.14 | 1.01 | 0.92 | 1.11 | 0.99 | 0.89 | 1.09 | 0.98 | 0.89 | 1.08 |
|  |  | High vs no exposure | | 0.84 | 0.66 | 1.07 | 1.04 | 0.88 | 1.21 | 1.11 | 0.92 | 1.35 | 1.04 | 0.89 | 1.20 |
| T3 | Q1: Most deprived | Low vs high | | 1.38 | 1.04 | 1.82 | 1.45 | 1.12 | 1.86 | 1.13 | 0.88 | 1.45 | 1.10 | 0.91 | 1.33 |
|  |  | Low vs no exposure | | 0.96 | 0.86 | 1.07 | 1.00 | 0.89 | 1.11 | 0.97 | 0.86 | 1.09 | 0.98 | 0.87 | 1.10 |
|  |  | High vs no exposure | | 0.70 | 0.53 | 0.92 | 0.69 | 0.54 | 0.88 | 0.86 | 0.67 | 1.10 | 0.89 | 0.74 | 1.07 |
|  | Q2 | Low vs high | | 1.30 | 1.00 | 1.67 | 1.24 | 0.99 | 1.55 | 1.09 | 0.85 | 1.39 | 0.99 | 0.82 | 1.19 |
|  |  | Low vs no exposure | | 1.02 | 0.92 | 1.13 | 1.00 | 0.90 | 1.11 | 0.87 | 0.78 | 0.98 | 0.86 | 0.77 | 0.96 |
|  |  | High vs no exposure | | 0.79 | 0.61 | 1.01 | 0.80 | 0.64 | 1.00 | 0.81 | 0.64 | 1.02 | 0.87 | 0.73 | 1.03 |
|  | Q3: Least deprived | Low vs high | | 1.26 | 1.01 | 1.57 | 1.32 | 1.08 | 1.61 | 1.03 | 0.84 | 1.28 | 1.07 | 0.92 | 1.25 |
|  |  | Low vs no exposure | | 1.01 | 0.92 | 1.12 | 1.02 | 0.93 | 1.12 | 0.96 | 0.87 | 1.07 | 1.01 | 0.91 | 1.11 |
|  |  | High vs no exposure | | 0.81 | 0.65 | 1.00 | 0.78 | 0.64 | 0.95 | 0.93 | 0.76 | 1.14 | 0.94 | 0.81 | 1.09 |

Note: All estimates from GEE models adjusted for maternal age, race/ethnicity, insurance payor, preexisting diabetes, chronic hypertension, obesity, chronic renal disease, abnormal presentation, prior cesarean, parity, placental abnormality, adequacy of prenatal care, smoking, year of birth, and maternal depression. Pre=Preconception, T1= Trimester 1, T2= Trimester 2, T3=Trimester 3,RR= Relative Risk, CI= 95% Confidence Interval, ICE: Index of Concentration of racialized, economic extremes. Q1: Low-income, majority Black communities compared to Q3: High-income, majority white communities.
